# Projected population health, demographics, and associated healthcare resource requirements from 2025 to 2047: modelling study in England

**DOI:** 10.64898/2025.12.22.25342817

**Authors:** Luke F Shaw, Zehra Onen-Dumlu, Christos Vasilakis, Richard M Wood

## Abstract

**Objectives:** There is a deficit of information available to guide longer-term healthcare planning in meeting health needs and ensuring financial sustainability. This study attempts to address this gap through projecting future population health state and demographics and associated healthcare resources required to satisfy expected demands over the next two decades.

**Methods:** A mathematical model is developed for projecting a population’s future age and health state, subject to births, deaths, immigration and emigration. These modelled outputs are combined with healthcare resource utilisation profiles to provide future cost estimates at a total level and for constituent healthcare settings. The model is calibrated to linked longitudinal patient-level data from a one-million-resident NHS healthcare system in England covering a mixture of urban, rural and coastal locations.

**Results:** Cost growth is projected to exceed population growth over the period from 2025 to 2047, with costs growing by 15-22%, depending on the scale of international migration, and the population growing by 4-16%. This is being driven by a projected 15-30% growth in the size of all but the healthiest population health states. If budgets were constrained to population growth, then either a 4% productivity improvement would be required or a 10% reduction in those transitioning to worse health states.

**Conclusions:** An ageing and unhealthier population is leading to a 5-11% rise in average per-person healthcare costs. To avert this, healthcare administrators may need to prioritise the quicker-acting cash-releasing productivity schemes to fund the prevention measures that may ultimately be required to secure longer-term financial sustainability.

## 1. Introduction

Effective planning of healthcare services requires an appreciation of the population’s healthcare needs both now and into the future, as well as an understanding of the value of interventions within the purview of healthcare administrators. For instance, estimates of the future size, age and health state of the population may guide effective decision-making in regard of capacity provision and allocation: doctors and nurses of the appropriate clinical specialties may be proactively trained to meet anticipated demands and substantial capital investments, such as building new hospitals, can be driven by the expected size, composition and needs of catchment populations (e.g., why build new maternity wards in areas where fertility rates are expected to decline?). Furthermore, estimates of the efficacy of productivity and prevention efforts may help influence the amount of effort and resources directed to each, in striking a suitable balance between the more immediate but bounded impacts of productivity initiatives, in making shorter-term cost savings, and the more gradual population health improvements resulting from disease prevention schemes, which may take longer to release savings but be ultimately essential to securing longer-term financial sustainability.

This paper concerns the projection of metrics of population health, demographics, and associated healthcare resource requirements for a large NHS healthcare system in England from 2025 to 2047. The first aim is to project, for each future year, the healthcare utilisation and costs, both at a total level and across the various healthcare settings (primary care, secondary care, mental health, etc.). This is based on estimates of the future number of inhabitants and their age and health state (representing the key determinants of healthcare needs [Navickas et al, 2016; Atella et al, 2019]) and is under the assumptions that there is no change to the extent that given healthcare needs are met, and that there are no new productivity and prevention interventions. The second aim is to project the possible impact of new productivity and prevention interventions through considering a range of scenarios in which incremental variations are made to the unit-costs of healthcare activity (representing productivity initiatives) and the probabilities which individuals transition between health states (representing prevention initiatives). All projections are generated from a novel mathematical modelling framework constructed for the purposes of this study and based on a local longitudinal patient-level linked dataset for the healthcare system’s entire adult population (with the exception of future trajectories for immigration, emigration and births, which are obtained from official estimates for the local area).

There are both academic and practical motivations to this study. From an academic perspective, there is a deficit within the current literature of studies addressing the abovementioned aims. Recent modelling studies have projected future patterns of major illness and multimorbidity [Kingston et al, 2018; Watt et al, 2023; Head et al, 2025] or examined long-term funding, beds, and workforce needs at a national level [Rachet-Jacquet et al, 2023]. These models typically focus on disease incidence, risk factors, or macro-level expenditure, instead of the provision of detailed projections of multi-setting healthcare utilisation and costs. Furthermore, these studies lack consideration to the individual and combined effects of productivity and prevention efforts on projected healthcare resource requirements. From a practical perspective, while healthcare planners appear to acknowledge the value in considering multiple years and decades ahead, there is a lack of evidenced progress in doing so within forward plans. For instance, only in 2025 has the NHS in England pledged to “shift from single to multi-year operational and financial planning” [NHS England, 2025]. And even the NHS’s Joint Forward Plan exercise, which requires assessments of “the local community’s current and future health, care and wellbeing needs to inform local decision-making and integrated care strategy”, covers just the next five years [NHS England, 2024]. Yet the directions outlined in the 2025-published NHS 10 Year Plan are heavily premised on the projected circumstances over a longer timeframe, with high-level estimates of rising disease prevalence to 2040 being cited as a key determinant to “the financial sustainability of the NHS” [UK Government, 2025]. The results of this study could help bridge this gap in providing useful methods and insights to support healthcare administrators in developing service plans of both sufficient detail and reach into the years and decades ahead.

## 2. Methods

### 2.1 Study setting and data

The studied healthcare system covers a one-million resident population in and around the major city of Bristol, in South West England. The population is demographically similar to England and is distributed over a mixture of urban, rural and coastal locations: Bristol is a major city with a relatively younger and diverse population; North Somerset contains a large coastal town and areas of severe deprivation; and South Gloucestershire covers a relatively affluent rural population. Together, these three geographies comprise the BNSSG healthcare system. Primary care is provided through 74 GP practices with secondary care delivered in three acute hospitals. There is a single community care provider and a single mental health provider. Telephone-based access to urgent care is through the 111 advice service and the 999 ambulance service. The planning and coordination of healthcare across these settings is the responsibility of the NHS BNSSG Integrated Care Board (ICB).

The local data for this study is provided by BNSSG ICB over the range from April 2021 to March 2024, i.e., the three financial years from 2021/22 to 2023/24. The System Wide Dataset is an existing resource that has, since 2019, linked patient-level data for primary care, secondary care, community care, mental health, 111 and 999 services for the BNSSG population (excluding the 1.7% of individuals who have opted out of sharing data) [BNSSG Integrated Care Board, 2025]. The two main tables within the System Wide Dataset are the Attributes and Activity tables. The Attributes table consists of a monthly snapshot of each person’s demographic, socioeconomic and clinical characteristics at that point in time. This includes variables such as age, sex and the presence of various chronic conditions. This data is sourced locally from the EMIS administration systems which are used by all GP practices within BNSSG. The Activity table contains information for all patient contacts over the abovementioned range of healthcare settings. This includes variables such as the date and time of the activity (whether it be a GP appointment, ED attendance, or community care visit), the specialty, provider and cost. With the exception of the primary care data, all other data is sourced from national commissioning dataflows provided to the ICB by NHS England. Data is linked across the two tables by the NHS number. All records within the System Wide Dataset use an anonymised version of the NHS number, with no storage of other person identifiable information (e.g., patient names or addresses). Use of the System Wide Dataset was approved for this study by the ICB’s cross-system data oversight group. Specific ethics approvals were not required due to the use of anonymised data.

The BNSSG healthcare system employs a segmentation approach to broadly categorise the health state of adult members of the population (17 years and older). Each person is assigned to one of five Core Segments (CS) according to their multimorbidity burden as measured through their Cambridge Multimorbidity Score (CMS) [Payne et al, 2020]. CMS is calculated by the weighted addition of 37 chronic conditions, whose prevalence is recorded in the Attributes table for each patient for each past month. CMS uniquely determines the CS based on four pre-fixed sequential thresholds, e.g., those with CMS < 0.09 (low multimorbidity) fall into CS1 (most healthy) and those with CMS ≥ 2.95 (high multimorbidity) fall into CS5 (least healthy). At the time at which the four thresholds were calibrated (in 2022), CS1 contained approximately half of the adult population and each sequentially higher-order Core Segment approximately halved in size and doubled in per-person spend. For more details on the Core Segment approach and its development, see [Wood et al, 2024]. In this current study, the Core Segments are used to represent each person’s health state.

Based on the latest available data for this study, from March 2024, we can report the age and Core Segment composition of the BNSSG adult population (Figure 1A), alongside the population pyramid (Figure 1B) and the geographical distribution of the population (Figure 1C).

**Figure 1.**
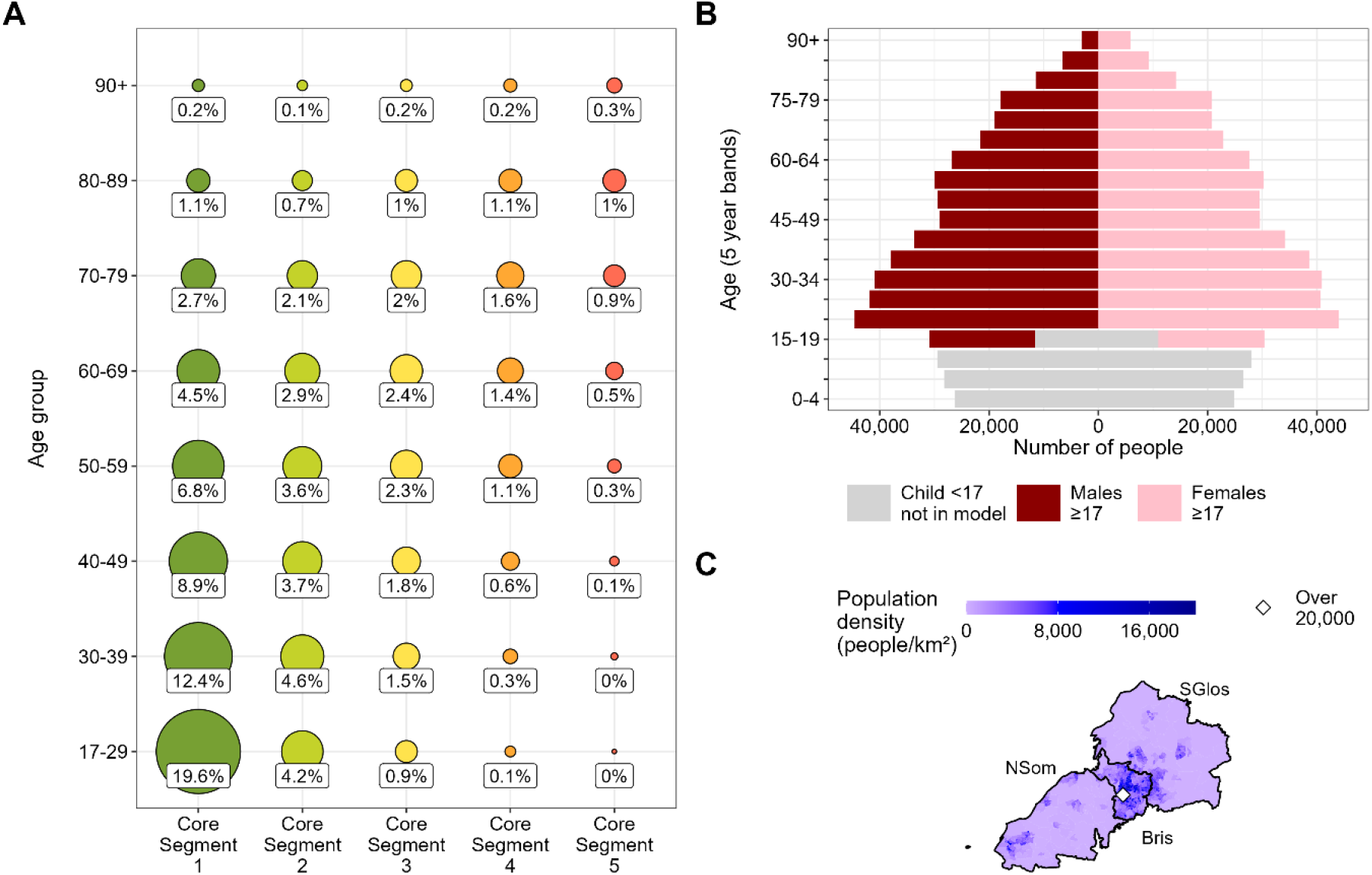
Summary of the studied population based on data from March 2024, consisting of [A] proportions of the population by health state (Core Segment) and age group, [B] age and sex profile, and [C] geographical distribution among the three constituent areas in BNSSG. Note: ‘Bris’ is Bristol; ‘NSom’ is North Somerset; and ‘SGlos’ is South Gloucestershire).

### 2.2 Model framework

As mentioned in Section 1, projected healthcare resource requirements are based on the projected number of inhabitants and their age and health state. As such, the modelling framework comprises two steps: the first is to annually project the number of people by age and Core Segment in each geography within the healthcare system, and the second is to convert these to healthcare utilisation and costs. This sub-section describes the model framework, with the input parameter values used in this study (as derived from local data for the BNSSG healthcare system) being covered in Section 2.3.

#### 2.2.1 Step 1: projecting the future population by age and health state

Step 1 is approached by defining the current composition of the adult population from which incremental perturbations are made in accounting for the amounts of ageing, health state changes, ‘births’ (local inhabitants reaching adulthood), deaths, immigrations and emigrations that occur within each future year. For each such future year, *y*, the composition of the population is measured by the number of individuals of a given age (in years), *a*, in a given Core Segment, *c*, and in a given geography within the healthcare system, *g*; which is represented by *n*_*y,g,a,c*_, where *y* ∈ *Y* = {0,1,2, …}, *g* ∈ *G* = {Bristol, North Somerset, South Gloucestershire}, *a* ∈ *A* = {17,18, …}, and *c* ∈ *C* = {1,2,4,4,5}. Thus, as of year *y*, the total population size is 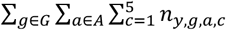 and – for example – the total number of individuals of age *a* is 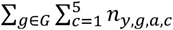.

Estimation of the *n*_*y,g,a,c*_ is as follows. Denoting the current year by *y* = 0, the current composition of the population is given by *n*_0,*g,a,c*_, which are assumed known. Projections for the following year (*y* = 1) are given by:

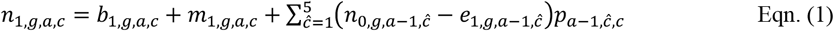

Here, *b*_1,*g,a,c*_ and *m*_1,*g,a,c*_ represent the number of local ‘births’ and new immigrants (from outside the geography), respectively, in year *y* = 1 for geography *g*, age *a* and Core Segment *c* (clearly, *b*_*y,g,a,c*_ = 0 for *a* ≠ 17). The right-most terms in Eqn. (1) represent the contribution from those individuals who were in the geography in year *y* = 0 and have not died nor emigrated and have since moved into the state representing age *a* and Core Segment *c*. From an age perspective, this cohort is restricted to those of age *a* − 1 in year *y* = 0 (since, of course, they will then be age *a* by year *y* = 1). From a Core Segment perspective, Core Segment *c* can be reached from any Core Segment *ĉ* ∈ *C* albeit with varying likelihoods. We represent by *p*_*a*−1,*ĉ,c*_ the probability of an individual of age *a* − 1 and Core Segment *ĉ* transitioning to Core Segment *c* in the next one year, assuming no emigration. We therefore subtract the emigration in year *y* = 1, *e*_1,*g,a*−1,_ *ĉ*, from the *n*_0,*g,a*−1,_ *ĉ* to determine the possible candidate number of individuals. To these we thereafter multiply through by *p*_*a*−1,_ *ĉ*_,*c*_ to restrict to the subset who transition to given Core Segment *c*. Note that we assume the *p*_*a*,_ *ĉ*_,*c*_ are calibrated such that 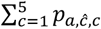 (i.e., the sum of one-year transition probabilities to any Core Segment) will not necessarily sum to one, with the remainder 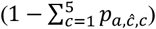 representing the one-year probability of death for those of age *a* and Core Segment *ĉ*.

Generalising, Eqn. (1) can be applied recursively for projecting into year *y* = 2 and, in turn, all subsequent considered years, and so we can write:

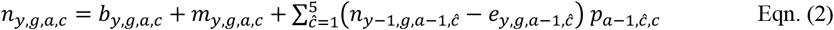

for all *y* ≥ 1.

#### 2.2.2 Step 2: projecting future healthcare resource requirements

Letting *τ*_*s,a,c*_ be the mean annualised healthcare resource cost for healthcare setting *s* ∈ *S* = {Primary care, Secondary care, etc.} for an individual of age *a* and Core Segment *c*, we estimate the total annual healthcare resource cost for year *y* for geography *g* for healthcare setting *s* for all individuals of age *a* and Core Segment *c* as:

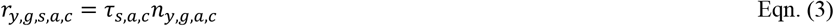

Hence, the estimated total resource cost for year *y* for healthcare setting *s* is 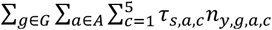 and the estimated overall total resource cost for year *y* is 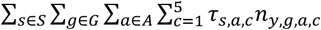.

Note that, within this formulation, no attempt is made to capture future inflation rates and so any future resource cost estimate is at current year prices. The reasons for this are, firstly, future inflation rates are difficult to predict and, secondly, including inflation would obscure the ability to clearly distinguish the effects of any future changes in population and health state.

### 2.3 Model calibration and scenario analysis

We now detail how the model described in Section 2.2 is fitted to the data described in Section 2.1. This requires the estimation of the following model parameters: *n*_1,*g,a,c*_, *p*_*a, ĉ,c*_, *b*_*y,g,a,c*_, *m*_*y,g,a,c*_, *e*_*y,g,a,c*_, and *τ*_*s,a,c*_. Note that a full specification of the model parameter values can be found in Section 1 of the Supplementary Material.

First consider the current composition of the population given by *n*_0,*g,a,c*_. These are based on local data for the numbers of the adult population in each geography *g* and of each age *a* and Core Segment *c* as of March 2024 (i.e., the latest month in the available data).

Second consider the Core Segment transition probabilities given by *p*_*a, ĉ,c*_. These are defined from the historical one-year volumes of Core Segment transitions as observed in local data from March 2021 to March 2024, excluding any within-one-year emigrations (but not deaths, for reasons mentioned in Section 2.2). Geography would not reasonably be expected to influence these parameters, and so data is aggregated over the considered geographies. The data is also aggregated to eight age groups, defined by *Ā* = {{17, …,29}, {40, …,49}, {40, …,49}, {50, …,59}, {60, …,69}, {70, …,79}, {80, …,89}, {90 +}}. These aggregations also help reduce the volatility associated with making estimates from smaller data subsets (Figure 1A). Let 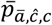 represent the resulting one-year transition probabilities, where *ā* ∈ *Ā* (Figure 2A). Thus 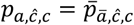 where *a* ∈ *ā*. E.g., the one-year transition probability from Core Segment *ĉ* to *c* for someone aged 25 is 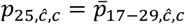.

**Figure 2.**
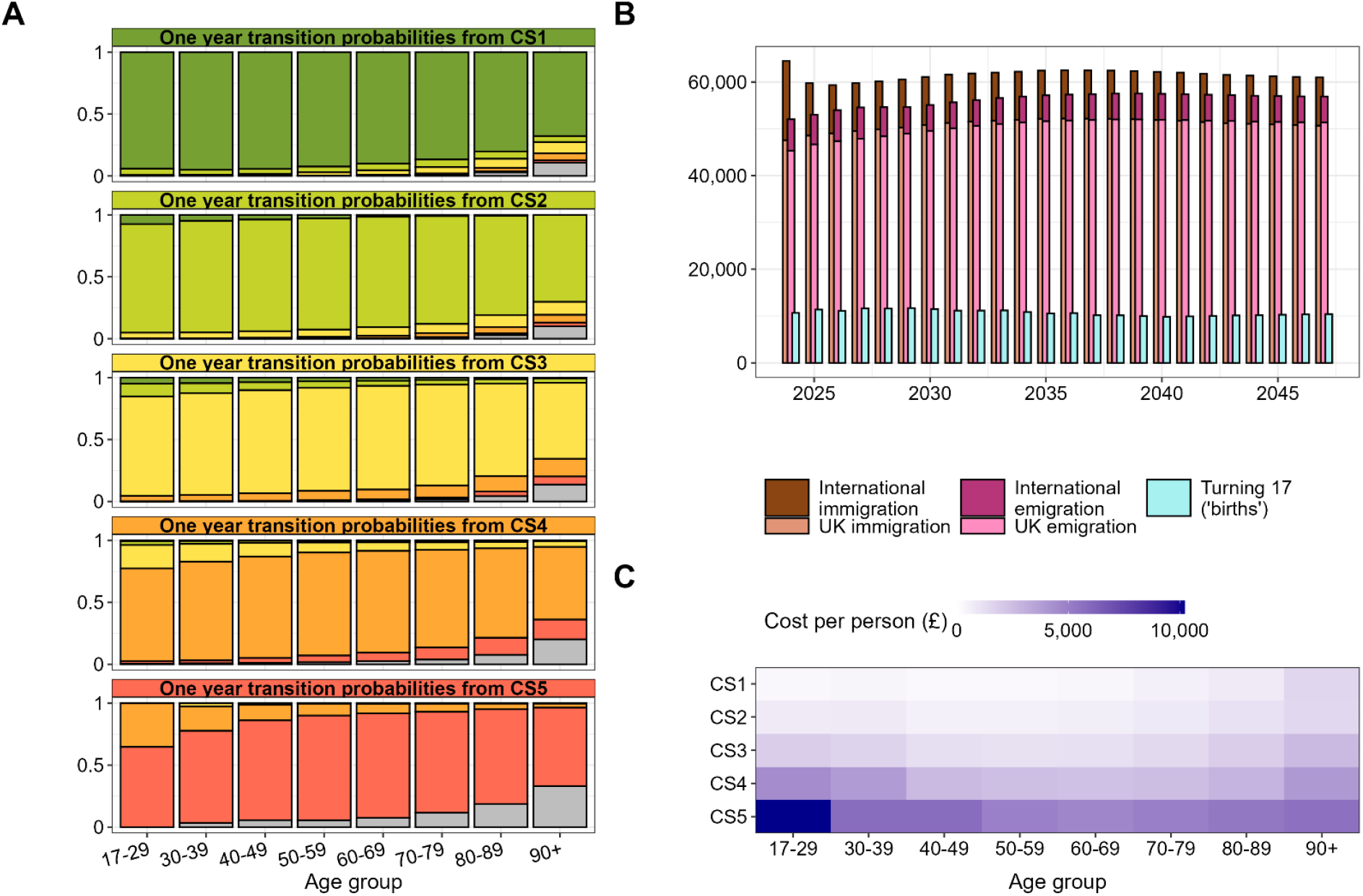
Summary of key parameters used within the modelling, consisting of [A] one-year transition probabilities between the health states (Core Segments) and to death (in grey), [B] annual rates of immigration and emigration (partitioned by whether the migration is international or to/from other parts of the UK) and ‘births’ (local inhabitants turning 17 years old), and [C] mean annual per-person healthcare costs by health state (Core Segment) and age group. Note: ‘CS’ is Core Segment.

Third consider the ‘birth’, immigration and emigration trajectories given by *b*_*y,g,a,c*_, *m*_*y,g,a,c*_, and *e*_*y,g,a,c*_ respectively. Official projections by the Office for National Statistics (ONS) are provided for each of these for each year until 2047, albeit, of course, with no Core Segment breakdown [Office for National Statistics, 2025]. These were published in June 2025 and are defined over a number of scenarios. Here, we consider a ‘Baseline lower’ and ‘Baseline upper’ scenario, based respectively on the ONS’s zero international migration scenario and its principal projection (Figure 2B). The reason for considering the zero international migration scenario is that the principal projection is based on actual data up to 2022, which covers a period of substantially elevated international immigration. The projections for 2023 and 2024 already considerably overstate the actual levels observed, and so, while it is recognised that international migration will not reach zero, this scenario offers a lower bound to the upper bound provided by the principal projection. Let the birth, immigration and emigration trajectories be represented by *B*_*y,w,g,a*_, *M*_*y,w,g,a*_, and *E*_*y,w,g,a*_, where scenario *w* ∈ *W* = {Baseline lower, Baseline upper} (of course, *B*_*y,w,g,a*_ = 0 for *a* ≠ 17). These are partitioned to Core Segment level by the historical proportions observed in local data from March 2021 to March 2024. Denote by 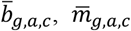, and 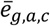 the observed proportions of total births, immigrations and emigrations in geography *g* in age *a* that are with respect to Core Segment *c*. I.e., 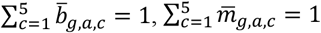, and 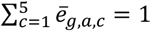 for any *g* and *a*. Thus, 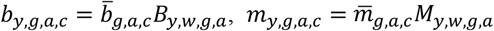, and 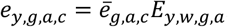.

Fourth consider the mean annualised healthcare resource costs given by *τ*_*s,a,c*_. The range of studied healthcare settings, *S*, covers primary care, secondary care, community care, mental health, 111 and 999 services. Each *τ*_*s,a,c*_ is based on mean costs observed in local data from March 2021 to March 2024 (totals over *S* displayed in Figure 2C). For similar reasons as before, the data is aggregated over the considered geographies and eight age groups *Ā*, resulting in 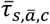. Thus 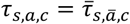 where *a* ∈ *ā*.

Finally, we make perturbations to the transition probabilities (*p*_*a, ĉ,c*_) and healthcare costs (*τ*_*s,a,c*_) in representing possible future initiatives relating to prevention and productivity. These are important in guiding economical use of limited resources as is mentioned prominently in the NHS 10 Year Plan [UK Government, 2025]. This talks of “a radical new path” for prevention with “bold action on obesity and tobacco”, as well as “boosting productivity significantly”. Accordingly, we consider a wide range of perturbations to the upward Core Segment transition probabilities (−70% to 70%) and mean healthcare resource costs (−25% to 50%), which also reflect the possibility of further deterioration. Furthermore, we consider two bespoke scenarios aimed at making specific improvements to cohorts potentially most amenable to intervention: we consider making a modest productivity improvement for the older and most costly cohort (5% reduced costs for people aged over 50 and in Core Segments 4 and 5) and improved disease prevention for the younger cohort (a 20% reduction in the upward transition probabilities for those under 40). The former is in line with UK Government plans (“for the next 3 years we have set the NHS a target to deliver a 2% year on year productivity gain” [UK Government, 2025]) and the latter is consistent with recent findings from disease reduction initiatives (such as a 20% lowering in the risk of developing Type 2 diabetes [Ravindrarajah et al, 2023]).

## 3. Results

The population is expected to grow by 16% to 2047 under the ‘Baseline upper’ birth and migration trajectories (Figure 3A), with 38% growth in those aged 80 and over, 18% growth in those aged between 30 and 79, and an initial 14% rise in those aged 17-29 tailing off to an eventual 5% increase on 2024 numbers (Figure 3B). While all Core Segments are larger, the least growth is in Core Segment 1 (representing the healthiest individuals), with 25-30% growth in all other Core Segments, indicating a progressively unhealthier population (Figure 3C). There is a considerable influence from international migration: using the birth and migration trajectories under the ‘Baseline lower’ scenario, the population would grow by only 4% to 2047, with substantially fewer numbers of younger and healthier individuals than under the ‘Baseline upper’ scenario.

**Figure 3.**
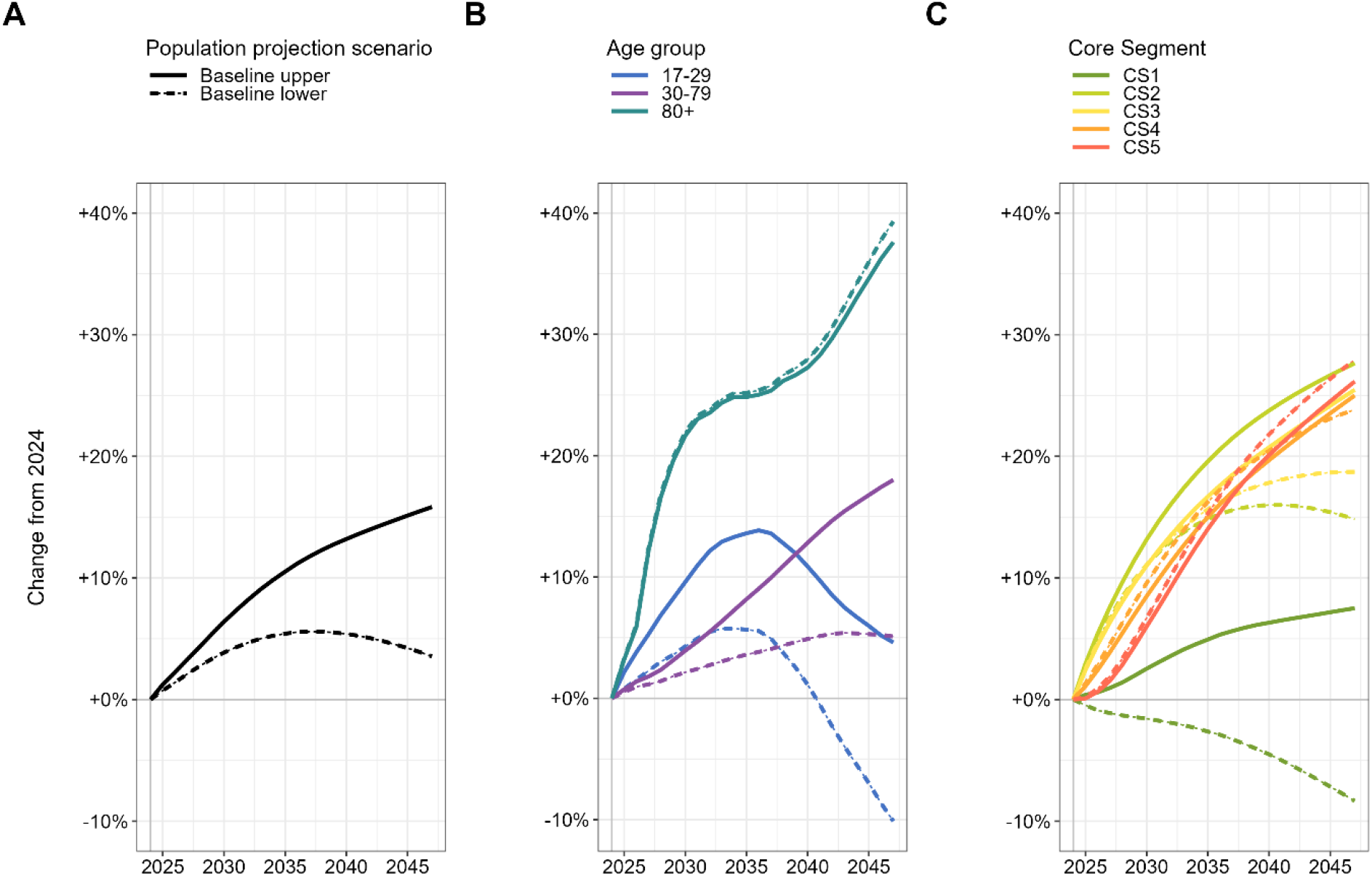
Projections, under the lower and upper Baseline scenarios, for [A] total population size and partitioned by [B] age and [C] health state (Core Segment).

Figure 4 shows how, under the ‘Baseline upper’ scenario, while the population is expected to grow by 16% (Figure 4A), the implications of an ageing and unhealthier population means that costs would grow at a disproportionate 22% (Figure 4B). Modelling also reveals the uneven geographical distribution of future changes, both in regard of the number of individuals (Figure 4A) and their healthcare requirements (Figure 4B). For instance, the large city of Bristol, with a greater proportion of relatively younger and healthier residents, is estimated to see costs rise in proportion to population growth (at 14%), whereas North Somerset, comprising more rural and coastal areas, is expected to see cost growth (28%) at more than double the population increase (13%). Fundamentally, this is being driven by an older population, which also explains the 44% growth in ‘Community’ healthcare resource costs in this geography (Figure 4C). As per the ‘Baseline lower’ scenario, international immigration has most impact on Bristol, with a 14 percentage point difference in population size with or without international migration. As would reasonably be expected, the younger profile of international immigrants means less (more) effect on healthcare settings typically serving older (younger) people: hence, the large differences for ‘Maternity’ costs and small differences for ‘Community’ costs.

**Figure 4.**
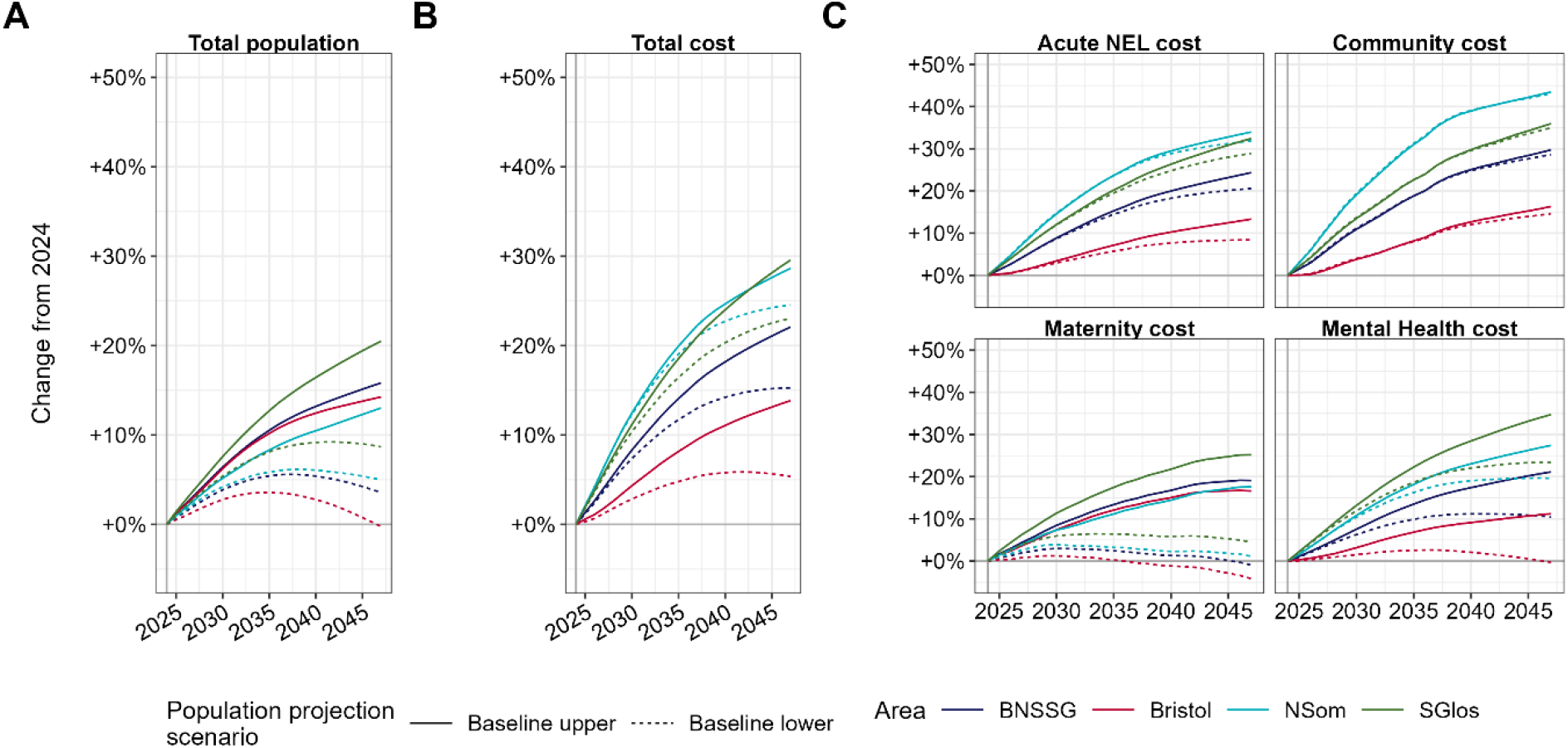
Projections, under the lower and upper Baseline scenarios, for [A] total population size and healthcare costs at a [B] total level and [C] partitioned by selected healthcare settings. Note: ‘NEL’ refers to non-elective admissions.

Figure 5A shows the range of impacts on total cost growth to 2047 from changes to healthcare unit-costs (representing productivity initiatives) and upward Core Segment transition probabilities (representing prevention initiatives), under the ‘Baseline upper’ scenario. The black square indicates the unadjusted ‘Baseline upper’ position, with the black solid line indicating the productivity/prevention trade-offs required to maintain total cost growth at 22%. So, for example, if shortfalls in workforce pushed up costs by 10%, then this would require the introduction of prevention efforts capable of reducing all upward Core Segment transitions by 20%, in order to maintain the equivalent 22% total cost growth to 2047. If healthcare budgets were increased only in line with population growth (16%), then either a 4% productivity increase has to be achieved or a 10% reduction in those transitioning to unhealthier health states (or combinations thereof as indicated by the dark grey solid line). A more targeted 5% productivity improvement to a costlier cohort – those aged over 50 in Core Segments 4 and 5 – would reduce total cost growth by 2 percentage points, with a greater reduction of 4 percentage points through 20% improved disease prevention for those aged under 40 (Figure 5B). The temporal cost profiles of these two examples illustrate the quicker but bounded impacts of the productivity initiative and the more gradual but sustained effects of the prevention initiative (Figure 5C). Ultimately, the more immediate marginal cost improvements of the productivity example are eclipsed by the prevention effort, as fewer numbers of those younger individuals whose disease has been prevented eventually end up in higher-order Core Segments in later life where costs are substantially greater.

**Figure 5.**
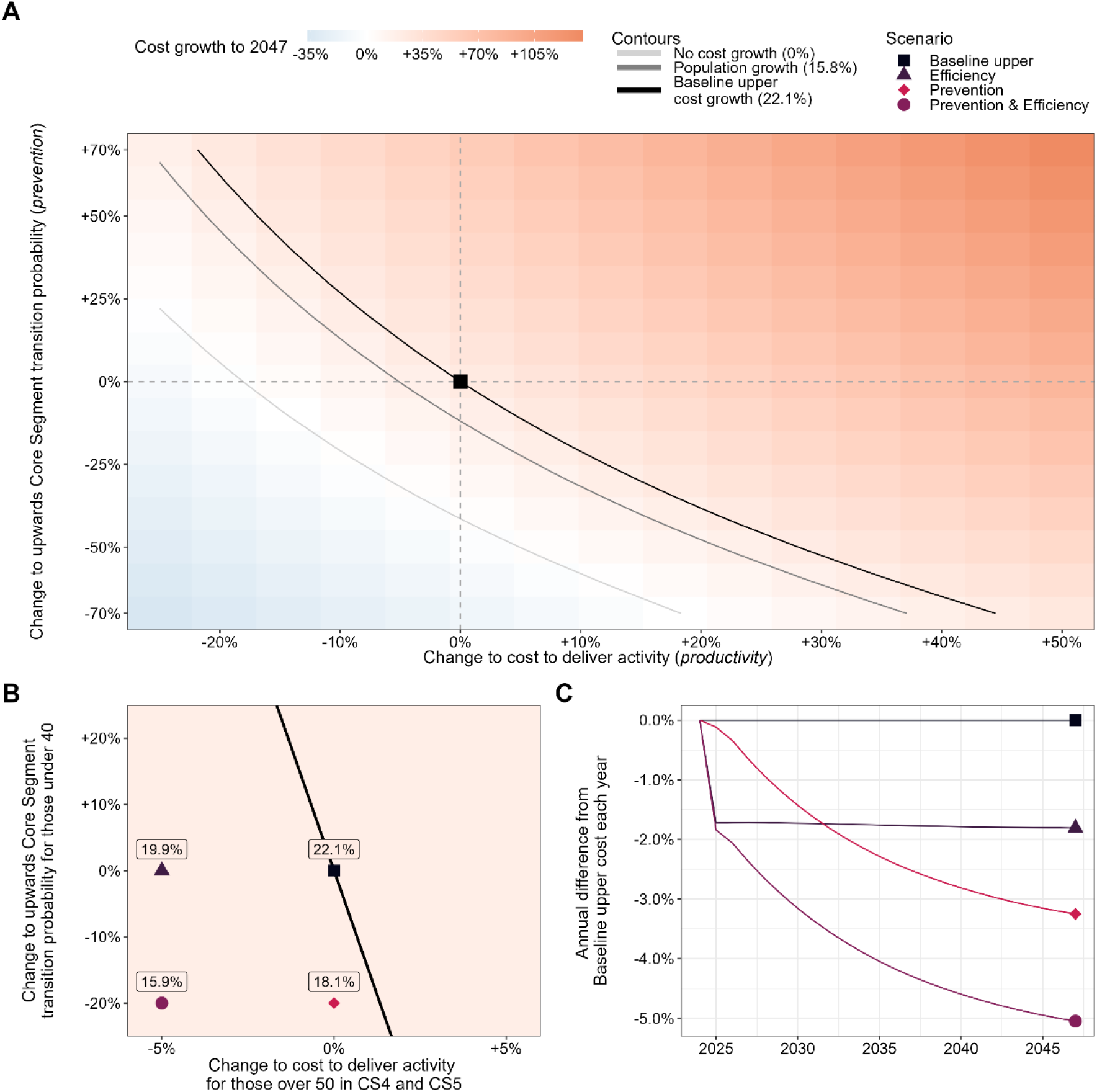
Scenario analysis for healthcare costs under the ‘Baseline upper’ scenario. [A] details the total cost growth from 2025 to 2047 according to variations on activity-level unit costs (representing productivity initiatives) and on upward Core Segment transition probabilities (representing prevention initiatives). The black square indicates the unadjusted ‘Baseline upper’ position with no productivity and prevention variations. Any point on the black solid line corresponds to the same total cost growth as the ‘Baseline upper’ scenario (22.1%); any point on the dark grey solid line corresponds to a total cost growth equivalent to the population growth over the period (15.8%); and any point on the light grey solid line corresponds to zero total cost growth. [B] and [C] consider a bespoke scenario aimed at making a modest productivity improvement for the older and most costly cohort (5% reduced costs for people aged over 50 and in Core Segments 4 and 5) and a bespoke scenario aimed at improving disease prevention for the younger cohort (a 20% reduction in the upward transition probabilities for those under 40), with [B] showing total cost growth from 2025 to 2047 and [C] showing the manifestation of these costs over time on comparison to the equivalent year of the ‘Baseline upper’ scenario.

Tabulated versions of the model results presented here can be found in Section 2 of the Supplementary Material.

## 4. Discussion

Recognising the current methodological and information deficits concerning longer-term health service planning, this study aimed to produce projections of population health, demographics, and associated healthcare resource requirements in England’s NHS from 2025 to 2047. Without intervention, this study estimates that cost growth would exceed population growth over this period, with costs growing by 15-22%, depending on the scale of international migration, and the population growing by 4-16% (Figure 4). This approximate 5-11% increase in average per-person healthcare costs is being driven by a progressively ageing and less healthy population – specifically, we project 15-30% growth in the size of all but the healthiest population health states (Figure 3C). This compares with an existing estimate cited in the NHS 10 Year Plan specifying a 37% increase in the number of people living with major illness by 2040 [Watt et al, 2023; UK Government, 2025]. Additionally, our study reveals the unequal distribution of projected demands, with particular pressure in both the geographic areas (rural locations) and healthcare settings (community care) most associated with older population cohorts (Figure 4C). A particular strength of this study is the use of a mature longitudinal patient-level linked dataset benefitting from both depth, in terms of primary care attribute data, and breadth, in terms of the considered range of healthcare settings. Another strength is the size of the studied healthcare system (covering a one-million resident population) and its representativeness of the England population as a whole (given demographic similarities and the mixture of urban, rural and coastal locations), thus supporting wider generalisability of the results.

The UK Government has documented its concerns regarding the financial sustainability of the NHS, stating that “if trends continue unimpeded … this would mean health spending rising at twice the real growth rate of the economy”, prompting the Prime Minister to declare “this is a time for radical change” [UK Government, 2025]. Appropriately – and representing a further strength to this study – our model is able to estimate the effects of hypothetical scenarios concerning productivity and prevention measures. This shows that, for cost growth to remain proportionate to population growth, a 4% productivity improvement or 10% reduction in those transitioning to worse health states would be required (Figure 5A). This could be achieved if the NHS delivered on the government’s productivity ambition: “for the next 3 years we have set the NHS a target to deliver a 2% year on year productivity gain” [UK Government, 2025]. The government is also committed to disease prevention, stating that “prevention is how we change this course”. Our modelling illustrates how prevention efforts, while slower acting, can bring about longer-lasting benefit (Figure 5C), and that – without any temporary financial headroom – healthcare administrators may need to urgently deliver the quicker cash-releasing productivity initiatives in order to fund the prevention measures that may be essential to ultimately securing longer-term financial sustainability.

At a local level, this modelling exercise has benefitted from close working with healthcare planners and senior management within the BNSSG system. Specifically, modelling insights have highlighted the need for versatility in considering the geographical distribution of future services and interventions, e.g., the need to develop alternative community-based services to absorb the otherwise-projected 30% increase in acute non-elective admission costs for the ageing North Somerset and South Gloucestershire populations (Figure 4C), if government plans to shift acute to community care are to be realised. Local healthcare managers have also signalled awareness of the need for agility given the substantial uncertainty in migration projections and the large consequential effects on the healthcare needs of the population (Figures 3 and 4) as well as implications for the size of the healthcare workforce required to meet those needs.

As with any empirically informed modelling study, there are a number of limitations to acknowledge. Fundamentally, this study assumes that health needs and corresponding healthcare demands can be adequately characterised through age and health state [Navickas et al, 2016; Atella et al, 2019], whereas existing knowledge suggests other factors such as sex and deprivation may also be contributory [Thompson et al, 2016; Collins et al, 2018]. In turn, we also assume that the five Core Segments offer sufficient discrimination in representing health state. Although empirically grounded, such a simplification neglects specific diseases and their particular impacts on a person’s health state and corresponding implications for health needs and healthcare demands. Relating to the Core Segments, the baseline scenarios assume the calibrated transition probabilities remain constant over the modelling horizon, whereas over time there would inevitably be changes in personal habits (e.g., diet and alcohol consumption) and clinical practice (e.g., new treatments for better managing risk factors). Also assumed constant are the rates of activity and costs overlaid to each age and Core Segment cohort, whereas realistically there would be changes in healthcare seeking behaviour (e.g., younger people seeking digital access routes [Lupton, 2021]) and the ways in which given health needs are met (e.g., improved technologies). Finally, our reliance on ONS demographic projections should be acknowledged – while these ‘official statistics’ are widely used for planning purposes, it is important to recognise the inherent uncertainty, which is somewhat addressed here through considering an upper and lower scenario on international migration.

There are various directions for future research in this domain. As well as better appreciating the range of variability in ONS projections, investigators could look to capture other sources of parameter and model uncertainty and assess their impacts on modelled outputs. Future work could also explore improvements to model sophistication, in regard of incorporating other appropriate determinants of health need and greater sensitivity in representing the presence and effect of certain diseases. As well as facilitating more robust parameter estimation, expanding the input data to include other geographic areas would improve representation, in ensuring wider coverage of other population cohorts and communities as well as other healthcare systems and operational practices.

## 5. Conclusions

An ageing and unhealthier population is leading to a projected 5-11% rise in average per-person healthcare costs. Projected additional resource requirements are unevenly distributed, with particular pressure in both the geographic areas (rural locations) and healthcare settings (community care) most associated with older population cohorts. To address these issues, healthcare administrators may need to prioritise the quicker-acting cash-releasing productivity schemes to fund the prevention measures that may ultimately be required to secure longer-term financial sustainability. Planners should also maintain awareness and agility to the healthcare resource implications of unknown future changes in population demographics and migration profiles.

## Data Availability

Data is not currently available due to legal restrictions on external sharing.

