## Supplementary Material for "Projected population health, demographics, and associated healthcare resource requirements from 2025 to 2047: modelling study in England"

Section SM.1 of this document complements Section 2.3 of the paper in providing the full specification of the parameters used to define the model.

Section SM.2 of this document complements Section 3 of the paper in providing tabulated versions of the model results.

### Section SM.1: Full specification of model parameters

#### *SM.1.1: Starting population, $n_{0,g,a,c}$*

*SM.1.1.1: For g = Bristol*

| Age | CS1 | CS2 | CS3 | CS4 | CS5 |
|---|---|---|---|---|---|
| 17 | 4564 | 446 | 80 | 2 | 0 |
| 18 | 4802 | 747 | 139 | 12 | 1 |
| 19 | 9357 | 1164 | 185 | 8 | 1 |
| 20 | 11251 | 1575 | 286 | 24 | 3 |
| 21 | 11173 | 1810 | 330 | 32 | 0 |
| 22 | 9105 | 1710 | 329 | 45 | 2 |
| 23 | 8367 | 1805 | 339 | 46 | 0 |
| 24 | 8324 | 1868 | 433 | 55 | 0 |
| 25 | 8078 | 1944 | 459 | 52 | 2 |
| 26 | 7240 | 2227 | 499 | 58 | 3 |
| 27 | 7500 | 2190 | 479 | 73 | 2 |
| 28 | 6941 | 2182 | 560 | 64 | 3 |
| 29 | 7040 | 2248 | 631 | 88 | 1 |
| 30 | 6632 | 2394 | 636 | 99 | 8 |
| 31 | 6073 | 2403 | 685 | 128 | 5 |
| 32 | 6083 | 2383 | 679 | 115 | 6 |
| 33 | 5942 | 2385 | 696 | 110 | 7 |
| 34 | 5724 | 2343 | 712 | 130 | 6 |
| 35 | 5499 | 2225 | 745 | 142 | 16 |
| 36 | 5059 | 2300 | 762 | 182 | 13 |
| 37 | 4952 | 2070 | 757 | 172 | 10 |
| 38 | 4874 | 2014 | 782 | 186 | 11 |
| 39 | 4610 | 1990 | 799 | 209 | 17 |
| 40 | 4179 | 1941 | 725 | 214 | 17 |
| 41 | 4088 | 1714 | 738 | 209 | 24 |
| 42 | 3989 | 1624 | 724 | 205 | 29 |
| 43 | 3701 | 1720 | 773 | 237 | 28 |
| 44 | 3780 | 1659 | 811 | 242 | 37 |
| 45 | 3457 | 1476 | 783 | 239 | 48 |
| 46 | 3112 | 1384 | 730 | 266 | 36 |
| 47 | 2931 | 1327 | 726 | 236 | 49 |
| 48 | 2944 | 1297 | 778 | 247 | 48 |
| 49 | 2777 | 1332 | 746 | 298 | 60 |
| 50 | 2614 | 1351 | 756 | 299 | 66 |
| 51 | 2449 | 1329 | 794 | 360 | 63 |
| 52 | 2361 | 1325 | 868 | 371 | 78 |
| 53 | 2523 | 1336 | 874 | 394 | 96 |
| 54 | 2250 | 1287 | 818 | 439 | 110 |
| 55 | 2173 | 1278 | 856 | 495 | 117 |
| 56 | 2241 | 1247 | 826 | 440 | 122 |
| 57 | 2138 | 1309 | 867 | 455 | 157 |
| 58 | 2106 | 1241 | 936 | 528 | 158 |
| 59 | 1804 | 1338 | 986 | 579 | 181 |
| 60 | 1715 | 1230 | 982 | 553 | 200 |
| 61 | 1598 | 1184 | 974 | 590 | 191 |
| 62 | 1452 | 1147 | 998 | 553 | 190 |
| 63 | 1441 | 1027 | 957 | 586 | 184 |
| 64 | 1260 | 993 | 878 | 541 | 196 |
| 65 | 1171 | 979 | 880 | 556 | 226 |
| 66 | 1110 | 958 | 867 | 575 | 247 |
| 67 | 1087 | 851 | 783 | 524 | 237 |
| 68 | 893 | 809 | 781 | 585 | 248 |
| 69 | 794 | 791 | 747 | 545 | 257 |
| 70 | 850 | 736 | 748 | 537 | 306 |
| 71 | 820 | 742 | 717 | 500 | 249 |
| 72 | 780 | 629 | 644 | 491 | 298 |
| 73 | 666 | 668 | 710 | 515 | 311 |
| 74 | 729 | 623 | 651 | 555 | 345 |
| 75 | 595 | 624 | 678 | 539 | 348 |
| 76 | 610 | 640 | 702 | 577 | 383 |
| 77 | 806 | 619 | 638 | 570 | 369 |
| 78 | 473 | 454 | 481 | 439 | 361 |
| 79 | 343 | 457 | 477 | 488 | 351 |
| 80 | 373 | 381 | 448 | 466 | 377 |
| 81 | 374 | 339 | 427 | 460 | 384 |
| 82 | 314 | 242 | 349 | 385 | 358 |
| 83 | 175 | 229 | 328 | 333 | 336 |
| 84 | 201 | 214 | 305 | 318 | 382 |
| 85 | 269 | 174 | 249 | 334 | 330 |
| 86 | 169 | 172 | 241 | 276 | 338 |
| 87 | 190 | 135 | 230 | 253 | 283 |
| 88 | 121 | 101 | 150 | 224 | 301 |
| 89 | 180 | 83 | 146 | 187 | 246 |
| 90 | 451 | 80 | 133 | 146 | 216 |
| 91 | 19 | 57 | 99 | 135 | 204 |
| 92 | 10 | 54 | 88 | 117 | 159 |
| 93 | 15 | 25 | 65 | 90 | 122 |
| 94 | 8 | 30 | 47 | 75 | 101 |
| 95 | 10 | 22 | 31 | 47 | 91 |
| 96 | 4 | 12 | 9 | 35 | 56 |
| 97 | 2 | 9 | 21 | 28 | 28 |
| 98 | 3 | 7 | 14 | 7 | 26 |
| 99 | 4 | 6 | 5 | 16 | 17 |

*SM.1.1.2: For g = North Somerset*

| Age | CS1 | CS2 | CS3 | CS4 | CS5 |
|---|---|---|---|---|---|
| 17 | 2299 | 170 | 32 | 3 | 0 |
| 18 | 2052 | 284 | 52 | 7 | 1 |
| 19 | 1263 | 321 | 63 | 3 | 2 |
| 20 | 1050 | 326 | 83 | 12 | 2 |
| 21 | 1220 | 313 | 75 | 12 | 0 |
| 22 | 1529 | 333 | 90 | 20 | 0 |
| 23 | 1678 | 409 | 99 | 9 | 0 |
| 24 | 1687 | 433 | 132 | 17 | 2 |
| 25 | 1697 | 488 | 141 | 18 | 0 |
| 26 | 1650 | 456 | 106 | 24 | 0 |
| 27 | 1526 | 527 | 151 | 22 | 1 |
| 28 | 1462 | 516 | 134 | 36 | 1 |
| 29 | 1489 | 543 | 157 | 36 | 1 |
| 30 | 1621 | 555 | 185 | 35 | 5 |
| 31 | 1769 | 560 | 159 | 38 | 1 |
| 32 | 1756 | 586 | 194 | 45 | 4 |
| 33 | 1883 | 578 | 208 | 42 | 1 |
| 34 | 1801 | 594 | 218 | 44 | 6 |
| 35 | 1784 | 631 | 234 | 69 | 7 |
| 36 | 1928 | 623 | 222 | 68 | 8 |
| 37 | 1923 | 603 | 212 | 68 | 4 |
| 38 | 1859 | 636 | 233 | 69 | 5 |
| 39 | 1844 | 614 | 244 | 87 | 7 |
| 40 | 1753 | 632 | 290 | 100 | 9 |
| 41 | 1846 | 627 | 289 | 80 | 9 |
| 42 | 1814 | 673 | 285 | 93 | 9 |
| 43 | 1861 | 647 | 308 | 107 | 11 |
| 44 | 1906 | 673 | 280 | 102 | 11 |
| 45 | 1830 | 599 | 290 | 116 | 10 |
| 46 | 1544 | 649 | 303 | 97 | 29 |
| 47 | 1583 | 608 | 316 | 139 | 22 |
| 48 | 1557 | 617 | 338 | 121 | 13 |
| 49 | 1671 | 616 | 355 | 143 | 19 |
| 50 | 1545 | 709 | 393 | 163 | 23 |
| 51 | 1574 | 702 | 380 | 196 | 22 |
| 52 | 1653 | 732 | 411 | 182 | 37 |
| 53 | 1709 | 760 | 437 | 206 | 39 |
| 54 | 1685 | 687 | 416 | 221 | 68 |
| 55 | 1646 | 766 | 450 | 218 | 52 |
| 56 | 1545 | 704 | 483 | 257 | 61 |
| 57 | 1614 | 714 | 517 | 244 | 71 |
| 58 | 1600 | 756 | 497 | 285 | 81 |
| 59 | 1583 | 737 | 529 | 268 | 96 |
| 60 | 1535 | 712 | 528 | 286 | 77 |
| 61 | 1465 | 739 | 530 | 289 | 82 |
| 62 | 1470 | 643 | 509 | 308 | 103 |
| 63 | 1372 | 638 | 468 | 266 | 105 |
| 64 | 1278 | 615 | 492 | 267 | 106 |
| 65 | 1131 | 604 | 547 | 284 | 110 |
| 66 | 1202 | 611 | 493 | 290 | 112 |
| 67 | 1081 | 538 | 458 | 287 | 132 |
| 68 | 1065 | 527 | 492 | 323 | 123 |
| 69 | 1054 | 494 | 442 | 297 | 148 |
| 70 | 954 | 521 | 478 | 332 | 141 |
| 71 | 1005 | 501 | 483 | 319 | 136 |
| 72 | 907 | 501 | 462 | 349 | 150 |
| 73 | 977 | 537 | 442 | 350 | 166 |
| 74 | 889 | 509 | 483 | 361 | 181 |
| 75 | 981 | 552 | 522 | 430 | 221 |
| 76 | 905 | 546 | 569 | 446 | 255 |
| 77 | 1021 | 604 | 581 | 482 | 300 |
| 78 | 822 | 393 | 454 | 385 | 233 |
| 79 | 682 | 422 | 467 | 382 | 302 |
| 80 | 766 | 361 | 364 | 345 | 306 |
| 81 | 548 | 311 | 373 | 318 | 258 |
| 82 | 505 | 275 | 293 | 308 | 216 |
| 83 | 420 | 210 | 258 | 272 | 254 |
| 84 | 328 | 173 | 243 | 288 | 279 |
| 85 | 435 | 157 | 218 | 243 | 264 |
| 86 | 292 | 145 | 223 | 218 | 228 |
| 87 | 297 | 132 | 181 | 186 | 227 |
| 88 | 182 | 115 | 156 | 176 | 224 |
| 89 | 217 | 75 | 137 | 161 | 209 |
| 90 | 542 | 56 | 101 | 135 | 179 |
| 91 | 13 | 60 | 84 | 93 | 155 |
| 92 | 15 | 53 | 77 | 96 | 137 |
| 93 | 10 | 30 | 50 | 76 | 112 |
| 94 | 9 | 31 | 34 | 62 | 92 |
| 95 | 7 | 15 | 33 | 37 | 73 |
| 96 | 8 | 17 | 25 | 34 | 52 |
| 97 | 1 | 10 | 13 | 12 | 33 |
| 98 | 2 | 10 | 11 | 12 | 23 |
| 99 | 1 | 10 | 8 | 10 | 12 |

*SM.1.1.3: For g = South Gloucestershire*

| Age | CS1 | CS2 | CS3 | CS4 | CS5 |
|---|---|---|---|---|---|
| 17 | 2299 | 170 | 32 | 3 | 0 |
| 18 | 2052 | 284 | 52 | 7 | 1 |
| 19 | 1263 | 321 | 63 | 3 | 2 |
| 20 | 1050 | 326 | 83 | 12 | 2 |
| 21 | 1220 | 313 | 75 | 12 | 0 |
| 22 | 1529 | 333 | 90 | 20 | 0 |
| 23 | 1678 | 409 | 99 | 9 | 0 |
| 24 | 1687 | 433 | 132 | 17 | 2 |
| 25 | 1697 | 488 | 141 | 18 | 0 |
| 26 | 1650 | 456 | 106 | 24 | 0 |
| 27 | 1526 | 527 | 151 | 22 | 1 |
| 28 | 1462 | 516 | 134 | 36 | 1 |
| 29 | 1489 | 543 | 157 | 36 | 1 |
| 30 | 1621 | 555 | 185 | 35 | 5 |
| 31 | 1769 | 560 | 159 | 38 | 1 |
| 32 | 1756 | 586 | 194 | 45 | 4 |
| 33 | 1883 | 578 | 208 | 42 | 1 |
| 34 | 1801 | 594 | 218 | 44 | 6 |
| 35 | 1784 | 631 | 234 | 69 | 7 |
| 36 | 1928 | 623 | 222 | 68 | 8 |
| 37 | 1923 | 603 | 212 | 68 | 4 |
| 38 | 1859 | 636 | 233 | 69 | 5 |
| 39 | 1844 | 614 | 244 | 87 | 7 |
| 40 | 1753 | 632 | 290 | 100 | 9 |
| 41 | 1846 | 627 | 289 | 80 | 9 |
| 42 | 1814 | 673 | 285 | 93 | 9 |
| 43 | 1861 | 647 | 308 | 107 | 11 |
| 44 | 1906 | 673 | 280 | 102 | 11 |
| 45 | 1830 | 599 | 290 | 116 | 10 |
| 46 | 1544 | 649 | 303 | 97 | 29 |
| 47 | 1583 | 608 | 316 | 139 | 22 |
| 48 | 1557 | 617 | 338 | 121 | 13 |
| 49 | 1671 | 616 | 355 | 143 | 19 |
| 50 | 1545 | 709 | 393 | 163 | 23 |
| 51 | 1574 | 702 | 380 | 196 | 22 |
| 52 | 1653 | 732 | 411 | 182 | 37 |
| 53 | 1709 | 760 | 437 | 206 | 39 |
| 54 | 1685 | 687 | 416 | 221 | 68 |
| 55 | 1646 | 766 | 450 | 218 | 52 |
| 56 | 1545 | 704 | 483 | 257 | 61 |
| 57 | 1614 | 714 | 517 | 244 | 71 |
| 58 | 1795 | 1115 | 734 | 316 | 61 |
| 59 | 1712 | 1028 | 750 | 352 | 89 |
| 60 | 1777 | 1064 | 748 | 304 | 79 |
| 61 | 1663 | 1012 | 683 | 350 | 103 |
| 62 | 1550 | 945 | 687 | 329 | 82 |
| 63 | 1342 | 888 | 706 | 335 | 123 |
| 64 | 1290 | 849 | 666 | 333 | 102 |
| 65 | 1137 | 821 | 654 | 359 | 97 |
| 66 | 1147 | 770 | 690 | 320 | 93 |
| 67 | 975 | 707 | 649 | 347 | 122 |
| 68 | 993 | 641 | 600 | 346 | 130 |
| 69 | 828 | 703 | 593 | 363 | 133 |
| 70 | 844 | 702 | 584 | 408 | 152 |
| 71 | 864 | 634 | 568 | 397 | 147 |
| 72 | 714 | 599 | 537 | 419 | 174 |
| 73 | 797 | 558 | 577 | 413 | 187 |
| 74 | 750 | 638 | 598 | 416 | 211 |
| 75 | 679 | 602 | 576 | 445 | 237 |
| 76 | 652 | 634 | 704 | 531 | 266 |
| 77 | 854 | 638 | 626 | 551 | 314 |
| 78 | 529 | 505 | 494 | 408 | 282 |
| 79 | 477 | 485 | 529 | 475 | 321 |
| 80 | 605 | 424 | 502 | 460 | 288 |
| 81 | 399 | 366 | 437 | 442 | 341 |
| 82 | 494 | 266 | 352 | 334 | 283 |
| 83 | 324 | 233 | 296 | 324 | 271 |
| 84 | 246 | 240 | 285 | 338 | 314 |
| 85 | 196 | 227 | 296 | 301 | 308 |
| 86 | 240 | 167 | 249 | 293 | 276 |
| 87 | 157 | 155 | 202 | 270 | 275 |
| 88 | 131 | 119 | 181 | 235 | 292 |
| 89 | 127 | 71 | 134 | 193 | 252 |
| 90 | 341 | 83 | 123 | 187 | 207 |
| 91 | 16 | 54 | 125 | 122 | 192 |
| 92 | 12 | 60 | 78 | 102 | 149 |
| 93 | 9 | 31 | 67 | 84 | 126 |
| 94 | 9 | 36 | 34 | 63 | 105 |
| 95 | 5 | 24 | 34 | 54 | 81 |
| 96 | 4 | 17 | 16 | 32 | 52 |
| 97 | 5 | 10 | 14 | 25 | 28 |
| 98 | 3 | 4 | 8 | 18 | 28 |
| 99 | 0 | 2 | 4 | 8 | 25 |

*SM.1.2: Core Segment transition probabilities,* $\overline{p}_{\overline{a},\hat{c},c}$

| Age group | From | To | | | | | |
|---|---|---|---|---|---|---|---|
| | | CS1 | CS2 | CS3 | CS4 | CS5 | Death |
| 17-29 | CS1 | 0.94137 | 0.05202 | 0.00627 | 0.00017 | 0 | 0.00018 |
| 17-29 | CS2 | 0.07369 | 0.87753 | 0.04589 | 0.00239 | 0 | 0.00051 |
| 17-29 | CS3 | 0.04773 | 0.10372 | 0.80346 | 0.04368 | 0 | 0.00141 |
| 17-29 | CS4 | 0.00571 | 0.03088 | 0.18831 | 0.75085 | 0.0188 | 0.00544 |
| 17-29 | CS5 | 0 | 0 | 0 | 0.35279 | 0.64721 | 0 |
| 30-39 | CS1 | 0.95097 | 0.03941 | 0.00913 | 0.00031 | 0 | 0.00019 |
| 30-39 | CS2 | 0.0479 | 0.90225 | 0.04562 | 0.00373 | 0 | 0.0005 |
| 30-39 | CS3 | 0.0447 | 0.08154 | 0.8203 | 0.0508 | 0.0008 | 0.00186 |
| 30-39 | CS4 | 0.00405 | 0.02338 | 0.14349 | 0.79745 | 0.02389 | 0.00774 |
| 30-39 | CS5 | 0 | 0 | 0.02518 | 0.19782 | 0.74455 | 0.03245 |
| 40-49 | CS1 | 0.94409 | 0.0394 | 0.01535 | 0.0007 | 0 | 0.00046 |
| 40-49 | CS2 | 0.03771 | 0.90143 | 0.05303 | 0.00664 | 0.0002 | 0.00099 |
| 40-49 | CS3 | 0.03667 | 0.06398 | 0.83273 | 0.06146 | 0.00168 | 0.00348 |
| 40-49 | CS4 | 0.00244 | 0.01679 | 0.11112 | 0.81905 | 0.03769 | 0.0129 |
| 40-49 | CS5 | 0 | 0 | 0.01189 | 0.12659 | 0.8058 | 0.05572 |
| 50-59 | CS1 | 0.92456 | 0.04689 | 0.02488 | 0.00228 | 0.00009 | 0.0013 |
| 50-59 | CS2 | 0.02736 | 0.89882 | 0.05902 | 0.01237 | 0.00051 | 0.00192 |
| 50-59 | CS3 | 0.0298 | 0.05056 | 0.83404 | 0.07617 | 0.0036 | 0.00583 |
| 50-59 | CS4 | 0.00218 | 0.014 | 0.08122 | 0.83213 | 0.05436 | 0.0161 |
| 50-59 | CS5 | 0 | 0 | 0.00414 | 0.09527 | 0.84604 | 0.05456 |
| 60-69 | CS1 | 0.90068 | 0.05328 | 0.03798 | 0.00476 | 0.0004 | 0.00289 |
| 60-69 | CS2 | 0.0157 | 0.89111 | 0.06829 | 0.01963 | 0.00136 | 0.00391 |
| 60-69 | CS3 | 0.02447 | 0.04143 | 0.83715 | 0.08145 | 0.00637 | 0.00913 |
| 60-69 | CS4 | 0.00159 | 0.01367 | 0.06915 | 0.82237 | 0.06827 | 0.02494 |
| 60-69 | CS5 | 0 | 0 | 0.00468 | 0.07751 | 0.84434 | 0.07346 |
| 70-79 | CS1 | 0.86735 | 0.06172 | 0.05127 | 0.01 | 0.00159 | 0.00807 |
| 70-79 | CS2 | 0.00977 | 0.87119 | 0.07519 | 0.03047 | 0.00407 | 0.0093 |
| 70-79 | CS3 | 0.01765 | 0.03898 | 0.81413 | 0.09699 | 0.01573 | 0.01652 |
| 70-79 | CS4 | 0.00135 | 0.01393 | 0.06098 | 0.78828 | 0.09786 | 0.0376 |
| 70-79 | CS5 | 0 | 0 | 0.00431 | 0.06492 | 0.81539 | 0.11538 |
| 80-89 | CS1 | 0.80156 | 0.05994 | 0.07397 | 0.02939 | 0.00742 | 0.02772 |
| 80-89 | CS2 | 0.00711 | 0.8012 | 0.09704 | 0.05198 | 0.01392 | 0.02876 |
| 80-89 | CS3 | 0.01228 | 0.03441 | 0.74852 | 0.12273 | 0.03944 | 0.04263 |
| 80-89 | CS4 | 0.00112 | 0.00928 | 0.05339 | 0.7219 | 0.13826 | 0.07605 |
| 80-89 | CS5 | 0 | 0 | 0.00345 | 0.04554 | 0.76381 | 0.1872 |
| 90+ | CS1 | 0.67799 | 0.04827 | 0.09134 | 0.05725 | 0.01782 | 0.10734 |
| 90+ | CS2 | 0 | 0.7023 | 0.10464 | 0.06299 | 0.02922 | 0.10085 |
| 90+ | CS3 | 0.00688 | 0.03414 | 0.61384 | 0.14324 | 0.0655 | 0.13639 |
| 90+ | CS4 | 0 | 0.00653 | 0.04534 | 0.58724 | 0.15973 | 0.20116 |
| 90+ | CS5 | 0 | 0 | 0.0034 | 0.03244 | 0.63366 | 0.3305 |

*SM.1.3: Projected births, $B_{y,w,g,a}$; immigrations, $M_{y,w,g,a}$; and emigrations, $E_{y,w,g,a}$*

Note the values reported here have been summed over all ages and are provided for the 'Baseline upper' ONS population projection.

| Year | Bristol | | | North Somerset | | | South Gloucestershire | | |
|---|---|---|---|---|---|---|---|---|---|
| | Births | Immigrations | Emigrations | Births | Immigrations | Emigrations | Births | Immigrations | Emigrations |
| 2024 | 4956 | 52044 | 44394 | 2493 | 11183 | 9365 | 3219 | 21632 | 17688 |
| 2025 | 5266 | 48736 | 45518 | 2648 | 11008 | 9447 | 3471 | 20856 | 18055 |
| 2026 | 5021 | 48421 | 46374 | 2606 | 11053 | 9554 | 3492 | 20864 | 18362 |
| 2027 | 5301 | 48819 | 46811 | 2768 | 11138 | 9657 | 3558 | 21049 | 18620 |
| 2028 | 5264 | 49080 | 46861 | 2796 | 11219 | 9704 | 3538 | 21210 | 18797 |
| 2029 | 5264 | 49404 | 46945 | 2769 | 11315 | 9765 | 3677 | 21387 | 18950 |
| 2030 | 5212 | 49834 | 47333 | 2650 | 11420 | 9850 | 3645 | 21596 | 19150 |
| 2031 | 5026 | 50208 | 47820 | 2589 | 11519 | 9922 | 3555 | 21782 | 19396 |
| 2032 | 4954 | 50389 | 48268 | 2613 | 11615 | 9967 | 3613 | 21913 | 19616 |
| 2033 | 4938 | 50484 | 48652 | 2669 | 11707 | 10009 | 3598 | 22005 | 19785 |
| 2034 | 4762 | 50602 | 48929 | 2551 | 11786 | 10072 | 3541 | 22090 | 19919 |
| 2035 | 4548 | 50749 | 49102 | 2455 | 11849 | 10139 | 3557 | 22174 | 20032 |
| 2036 | 4568 | 50728 | 49196 | 2523 | 11901 | 10166 | 3544 | 22218 | 20140 |
| 2037 | 4385 | 50631 | 49262 | 2420 | 11949 | 10171 | 3367 | 22235 | 20218 |
| 2038 | 4336 | 50533 | 49316 | 2406 | 11994 | 10204 | 3402 | 22247 | 20271 |
| 2039 | 4277 | 50389 | 49294 | 2357 | 12027 | 10208 | 3357 | 22229 | 20295 |
| 2040 | 4245 | 50160 | 49217 | 2279 | 12050 | 10214 | 3318 | 22190 | 20304 |
| 2041 | 4280 | 49973 | 49112 | 2295 | 12070 | 10209 | 3353 | 22154 | 20296 |
| 2042 | 4333 | 49628 | 48984 | 2309 | 12082 | 10196 | 3395 | 22084 | 20292 |
| 2043 | 4370 | 49356 | 48828 | 2323 | 12091 | 10198 | 3435 | 22012 | 20287 |
| 2044 | 4384 | 49179 | 48675 | 2329 | 12098 | 10210 | 3461 | 21968 | 20282 |
| 2045 | 4430 | 49011 | 48521 | 2344 | 12103 | 10225 | 3508 | 21927 | 20300 |
| 2046 | 4458 | 48845 | 48389 | 2359 | 12111 | 10239 | 3537 | 21893 | 20327 |
| 2047 | 4488 | 48708 | 48310 | 2372 | 12124 | 10255 | 3567 | 21867 | 20359 |

*SM.1.4: Core Segment breakdown for births, $\bar{b}_{g,a,c}$; immigrations, $\bar{m}_{g,a,c}$; and emigrations, $\bar{e}_{g,a,c}$*

*SM.1.4.1: For g = Bristol*

| Age | Births | | | | |
|---|---|---|---|---|---|
| | CS1 | CS2 | CS3 | CS4 | CS5 |
| 17 | 0.89001 | 0.09313 | 0.01645 | 0.00041 | 0 |

| Age | Immigrations | | | | | Emigrations | | | | |
|---|---|---|---|---|---|---|---|---|---|---|
| | CS1 | CS2 | CS3 | CS4 | CS5 | CS1 | CS2 | CS3 | CS4 | CS5 |
| 17 | 0.94107 | 0.05199 | 0.00347 | 0.00347 | 0.00000 | 0.88245 | 0.10145 | 0.01449 | 0.00161 | 0.00000 |
| 18 | 0.89052 | 0.09267 | 0.01681 | 0.00000 | 0.00000 | 0.85776 | 0.12508 | 0.01594 | 0.00123 | 0.00000 |
| 19 | 0.85727 | 0.12416 | 0.01831 | 0.00026 | 0.00000 | 0.79291 | 0.17661 | 0.02735 | 0.00313 | 0.00000 |
| 20 | 0.83363 | 0.14487 | 0.02025 | 0.00126 | 0.00000 | 0.78132 | 0.18821 | 0.03047 | 0.00000 | 0.00000 |

| | | | | | | | | | | |
|---|---|---|---|---|---|---|---|---|---|---|
| 21 | 0.83713 | 0.14076 | 0.01941 | 0.00270 | 0.00000 | 0.77047 | 0.19105 | 0.03545 | 0.00303 | 0.00000 |
| 22 | 0.85977 | 0.11460 | 0.02307 | 0.00211 | 0.00045 | 0.76137 | 0.19748 | 0.03863 | 0.00252 | 0.00000 |
| 23 | 0.84623 | 0.13058 | 0.02162 | 0.00156 | 0.00000 | 0.76867 | 0.18735 | 0.03997 | 0.00401 | 0.00000 |
| 24 | 0.81032 | 0.15567 | 0.03266 | 0.00135 | 0.00000 | 0.80107 | 0.16886 | 0.02627 | 0.00381 | 0.00000 |
| 25 | 0.80457 | 0.15964 | 0.03230 | 0.00350 | 0.00000 | 0.75122 | 0.20299 | 0.03820 | 0.00637 | 0.00122 |
| 26 | 0.77505 | 0.19201 | 0.03060 | 0.00234 | 0.00000 | 0.68601 | 0.25879 | 0.05032 | 0.00488 | 0.00000 |
| 27 | 0.79038 | 0.16176 | 0.04287 | 0.00499 | 0.00000 | 0.72938 | 0.20762 | 0.05670 | 0.00630 | 0.00000 |
| 28 | 0.78126 | 0.17574 | 0.04035 | 0.00265 | 0.00000 | 0.69892 | 0.24045 | 0.05217 | 0.00758 | 0.00087 |
| 29 | 0.76983 | 0.17626 | 0.04804 | 0.00587 | 0.00000 | 0.67216 | 0.25855 | 0.06044 | 0.00794 | 0.00092 |
| 30 | 0.75927 | 0.17527 | 0.05300 | 0.01178 | 0.00069 | 0.69435 | 0.23936 | 0.06141 | 0.00419 | 0.00070 |
| 31 | 0.77712 | 0.17623 | 0.04073 | 0.00592 | 0.00000 | 0.66116 | 0.25525 | 0.07602 | 0.00757 | 0.00000 |
| 32 | 0.78182 | 0.16996 | 0.03794 | 0.01028 | 0.00000 | 0.66910 | 0.24348 | 0.07055 | 0.01610 | 0.00077 |
| 33 | 0.77048 | 0.17278 | 0.05205 | 0.00341 | 0.00128 | 0.66449 | 0.24234 | 0.08092 | 0.01022 | 0.00204 |
| 34 | 0.73173 | 0.19380 | 0.06660 | 0.00694 | 0.00093 | 0.67136 | 0.22481 | 0.08887 | 0.01496 | 0.00000 |
| 35 | 0.78727 | 0.15380 | 0.04478 | 0.01296 | 0.00118 | 0.63868 | 0.25000 | 0.09069 | 0.02063 | 0.00000 |
| 36 | 0.76135 | 0.17250 | 0.05577 | 0.01038 | 0.00000 | 0.65162 | 0.24695 | 0.08178 | 0.01859 | 0.00106 |
| 37 | 0.76383 | 0.15461 | 0.05603 | 0.02553 | 0.00000 | 0.64646 | 0.24703 | 0.09065 | 0.01586 | 0.00000 |
| 38 | 0.75357 | 0.16244 | 0.06101 | 0.01981 | 0.00317 | 0.64421 | 0.24496 | 0.08753 | 0.01952 | 0.00378 |
| 39 | 0.73374 | 0.16219 | 0.08239 | 0.02168 | 0.00000 | 0.61353 | 0.26617 | 0.08947 | 0.02932 | 0.00150 |
| 40 | 0.73273 | 0.18233 | 0.05663 | 0.02605 | 0.00227 | 0.66641 | 0.23398 | 0.08031 | 0.01776 | 0.00154 |
| 41 | 0.76202 | 0.15290 | 0.06782 | 0.01480 | 0.00247 | 0.60757 | 0.26500 | 0.10342 | 0.02124 | 0.00277 |
| 42 | 0.74609 | 0.15885 | 0.06120 | 0.02865 | 0.00521 | 0.60852 | 0.25173 | 0.11397 | 0.02180 | 0.00396 |
| 43 | 0.70448 | 0.13585 | 0.11485 | 0.03641 | 0.00840 | 0.57998 | 0.25387 | 0.13106 | 0.03302 | 0.00206 |
| 44 | 0.74091 | 0.14242 | 0.08788 | 0.02576 | 0.00303 | 0.59422 | 0.26508 | 0.09925 | 0.03518 | 0.00628 |
| 45 | 0.70076 | 0.14583 | 0.11932 | 0.02652 | 0.00758 | 0.57649 | 0.24363 | 0.12748 | 0.03824 | 0.01416 |
| 46 | 0.66595 | 0.18415 | 0.10921 | 0.03640 | 0.00428 | 0.57183 | 0.24751 | 0.11664 | 0.05690 | 0.00711 |
| 47 | 0.72955 | 0.15000 | 0.06364 | 0.04545 | 0.01136 | 0.57076 | 0.20684 | 0.14774 | 0.06687 | 0.00778 |
| 48 | 0.72059 | 0.12745 | 0.11765 | 0.02451 | 0.00980 | 0.53541 | 0.24352 | 0.17271 | 0.03800 | 0.01036 |
| 49 | 0.66324 | 0.16710 | 0.12853 | 0.03085 | 0.01028 | 0.50090 | 0.27848 | 0.14105 | 0.06691 | 0.01266 |
| 50 | 0.67723 | 0.17291 | 0.09798 | 0.04035 | 0.01153 | 0.52688 | 0.27419 | 0.12903 | 0.05735 | 0.01254 |
| 51 | 0.69091 | 0.16970 | 0.09091 | 0.02727 | 0.02121 | 0.53271 | 0.24112 | 0.16075 | 0.05981 | 0.00561 |
| 52 | 0.66279 | 0.13372 | 0.11047 | 0.08721 | 0.00581 | 0.48545 | 0.27636 | 0.16182 | 0.06182 | 0.01455 |
| 53 | 0.67576 | 0.13939 | 0.13636 | 0.03030 | 0.01818 | 0.48771 | 0.28166 | 0.14178 | 0.07750 | 0.01134 |
| 54 | 0.62102 | 0.18790 | 0.12420 | 0.04777 | 0.01911 | 0.45800 | 0.22200 | 0.19200 | 0.10600 | 0.02200 |
| 55 | 0.67399 | 0.19780 | 0.06227 | 0.05128 | 0.01465 | 0.50209 | 0.26987 | 0.13808 | 0.06276 | 0.02720 |
| 56 | 0.62409 | 0.15693 | 0.17518 | 0.03650 | 0.00730 | 0.47454 | 0.25231 | 0.17824 | 0.07639 | 0.01852 |
| 57 | 0.61154 | 0.16923 | 0.14615 | 0.03462 | 0.03846 | 0.47309 | 0.24888 | 0.17937 | 0.07399 | 0.02466 |
| 58 | 0.63071 | 0.15353 | 0.11203 | 0.07884 | 0.02490 | 0.38600 | 0.29797 | 0.17833 | 0.12190 | 0.01580 |
| 59 | 0.52893 | 0.16529 | 0.16529 | 0.10331 | 0.03719 | 0.42069 | 0.27356 | 0.19770 | 0.08736 | 0.02069 |
| 60 | 0.50242 | 0.23188 | 0.16425 | 0.06763 | 0.03382 | 0.40988 | 0.28395 | 0.17778 | 0.09383 | 0.03457 |
| 61 | 0.55941 | 0.16832 | 0.15347 | 0.06436 | 0.05446 | 0.42697 | 0.23315 | 0.17416 | 0.10674 | 0.05899 |
| 62 | 0.61856 | 0.12887 | 0.13918 | 0.06186 | 0.05155 | 0.39130 | 0.26667 | 0.20000 | 0.11594 | 0.02609 |
| 63 | 0.54386 | 0.19298 | 0.18129 | 0.05263 | 0.02924 | 0.34347 | 0.27964 | 0.20669 | 0.12766 | 0.04255 |
| 64 | 0.54839 | 0.14839 | 0.19355 | 0.07742 | 0.03226 | 0.38000 | 0.25333 | 0.20000 | 0.12333 | 0.04333 |
| 65 | 0.52899 | 0.18116 | 0.15217 | 0.09420 | 0.04348 | 0.35827 | 0.28346 | 0.22047 | 0.09449 | 0.04331 |
| 66 | 0.48872 | 0.30075 | 0.13534 | 0.06015 | 0.01504 | 0.33871 | 0.26210 | 0.21371 | 0.12903 | 0.05645 |
| 67 | 0.46903 | 0.20354 | 0.15929 | 0.07965 | 0.08850 | 0.27982 | 0.33028 | 0.17431 | 0.12385 | 0.09174 |

| | | | | | | | | | | |
|---|---|---|---|---|---|---|---|---|---|---|
| 68 | 0.59048 | 0.19048 | 0.07619 | 0.05714 | 0.08571 | 0.34722 | 0.24537 | 0.22222 | 0.14815 | 0.03704 |
| 69 | 0.51064 | 0.14894 | 0.15957 | 0.12766 | 0.05319 | 0.29775 | 0.23034 | 0.22472 | 0.17416 | 0.07303 |
| 70 | 0.46067 | 0.17978 | 0.17978 | 0.13483 | 0.04494 | 0.33537 | 0.24390 | 0.21951 | 0.11585 | 0.08537 |
| 71 | 0.47619 | 0.13095 | 0.26190 | 0.08333 | 0.04762 | 0.24060 | 0.24812 | 0.27068 | 0.18045 | 0.06015 |
| 72 | 0.53947 | 0.18421 | 0.06579 | 0.10526 | 0.10526 | 0.25166 | 0.25166 | 0.26490 | 0.13907 | 0.09272 |
| 73 | 0.54167 | 0.11111 | 0.22222 | 0.05556 | 0.06944 | 0.24427 | 0.22901 | 0.25954 | 0.16794 | 0.09924 |
| 74 | 0.51429 | 0.11429 | 0.08571 | 0.14286 | 0.14286 | 0.25191 | 0.20611 | 0.23664 | 0.22137 | 0.08397 |
| 75 | 0.44444 | 0.05556 | 0.20833 | 0.09722 | 0.19444 | 0.25806 | 0.24194 | 0.20161 | 0.20161 | 0.09677 |
| 76 | 0.45205 | 0.12329 | 0.15068 | 0.20548 | 0.06849 | 0.20161 | 0.24194 | 0.21774 | 0.20161 | 0.13710 |
| 77 | 0.37662 | 0.09091 | 0.12987 | 0.22078 | 0.18182 | 0.16000 | 0.29000 | 0.20000 | 0.19000 | 0.16000 |
| 78 | 0.34483 | 0.18966 | 0.18966 | 0.08621 | 0.18966 | 0.17925 | 0.25472 | 0.23585 | 0.16038 | 0.16981 |
| 79 | 0.35714 | 0.10714 | 0.14286 | 0.17857 | 0.21429 | 0.18987 | 0.16456 | 0.29114 | 0.21519 | 0.13924 |
| 80 | 0.42188 | 0.20313 | 0.07813 | 0.17188 | 0.12500 | 0.08333 | 0.22619 | 0.20238 | 0.27381 | 0.21429 |
| 81 | 0.36207 | 0.15517 | 0.12069 | 0.15517 | 0.20690 | 0.12500 | 0.20833 | 0.29167 | 0.22222 | 0.15278 |
| 82 | 0.07692 | 0.11538 | 0.15385 | 0.26923 | 0.38462 | 0.14545 | 0.18182 | 0.21818 | 0.21818 | 0.23636 |
| 83 | 0.25532 | 0.17021 | 0.23404 | 0.14894 | 0.19149 | 0.09589 | 0.19178 | 0.21918 | 0.26027 | 0.23288 |
| 84 | 0.16000 | 0.04000 | 0.08000 | 0.36000 | 0.36000 | 0.10145 | 0.10145 | 0.27536 | 0.28986 | 0.23188 |
| 85 | 0.30909 | 0.10909 | 0.10909 | 0.16364 | 0.30909 | 0.07407 | 0.16667 | 0.16667 | 0.22222 | 0.37037 |
| 86 | 0.40385 | 0.00000 | 0.03846 | 0.15385 | 0.40385 | 0.04545 | 0.13636 | 0.34848 | 0.22727 | 0.24242 |
| 87 | 0.29630 | 0.11111 | 0.11111 | 0.18519 | 0.29630 | 0.07692 | 0.17308 | 0.17308 | 0.25000 | 0.32692 |
| 88 | 0.15385 | 0.11538 | 0.19231 | 0.15385 | 0.38462 | 0.05172 | 0.08621 | 0.29310 | 0.27586 | 0.29310 |
| 89 | 0.16667 | 0.06250 | 0.22917 | 0.27083 | 0.27083 | 0.12069 | 0.10345 | 0.13793 | 0.29310 | 0.34483 |
| 90+ | 0.23881 | 0.18905 | 0.18905 | 0.00000 | 0.38308 | 0.04790 | 0.07984 | 0.18962 | 0.30140 | 0.38124 |

*SM.1.4.2: For g = North Somerset*

| Age | Births | | | | |
|---|---|---|---|---|---|
| | **CS1** | **CS2** | **CS3** | **CS4** | **CS5** |
| 17 | 0.89784 | 0.08505 | 0.0156 | 0.00151 | 0 |

| Age | Immigrations | | | | | Emigrations | | | | |
|---|---|---|---|---|---|---|---|---|---|---|
| | **CS1** | **CS2** | **CS3** | **CS4** | **CS5** | **CS1** | **CS2** | **CS3** | **CS4** | **CS5** |
| 17 | 0.97354 | 0.01323 | 0.01323 | 0.00000 | 0.00000 | 0.88793 | 0.09806 | 0.01401 | 0.00000 | 0.00000 |
| 18 | 0.91190 | 0.07551 | 0.00000 | 0.01259 | 0.00000 | 0.85970 | 0.12276 | 0.01754 | 0.00000 | 0.00000 |
| 19 | 0.75266 | 0.22466 | 0.01134 | 0.00000 | 0.01134 | 0.75141 | 0.18796 | 0.05459 | 0.00604 | 0.00000 |
| 20 | 0.66965 | 0.25883 | 0.07152 | 0.00000 | 0.00000 | 0.71272 | 0.20804 | 0.07923 | 0.00000 | 0.00000 |
| 21 | 0.76572 | 0.19528 | 0.03900 | 0.00000 | 0.00000 | 0.66678 | 0.22226 | 0.10253 | 0.00843 | 0.00000 |
| 22 | 0.72773 | 0.23560 | 0.03667 | 0.00000 | 0.00000 | 0.67436 | 0.24993 | 0.07571 | 0.00000 | 0.00000 |
| 23 | 0.61954 | 0.33182 | 0.04864 | 0.00000 | 0.00000 | 0.60305 | 0.31298 | 0.06870 | 0.01527 | 0.00000 |
| 24 | 0.67131 | 0.20546 | 0.10501 | 0.01822 | 0.00000 | 0.68188 | 0.25312 | 0.05200 | 0.01300 | 0.00000 |
| 25 | 0.71229 | 0.21467 | 0.06829 | 0.00476 | 0.00000 | 0.64540 | 0.29080 | 0.04969 | 0.01411 | 0.00000 |
| 26 | 0.71883 | 0.23210 | 0.03581 | 0.01326 | 0.00000 | 0.59847 | 0.31380 | 0.08044 | 0.00728 | 0.00000 |
| 27 | 0.76261 | 0.18727 | 0.03657 | 0.01355 | 0.00000 | 0.65554 | 0.25695 | 0.07416 | 0.00667 | 0.00667 |
| 28 | 0.69960 | 0.21509 | 0.07182 | 0.00911 | 0.00437 | 0.58191 | 0.31337 | 0.08965 | 0.01508 | 0.00000 |
| 29 | 0.74578 | 0.20008 | 0.04570 | 0.00845 | 0.00000 | 0.60686 | 0.24670 | 0.12665 | 0.01319 | 0.00660 |
| 30 | 0.70195 | 0.22057 | 0.06528 | 0.00824 | 0.00396 | 0.67729 | 0.20000 | 0.09694 | 0.02576 | 0.00000 |

| | | | | | | | | | | |
|---|---|---|---|---|---|---|---|---|---|---|
| 31 | 0.70649 | 0.18227 | 0.10226 | 0.00897 | 0.00000 | 0.64566 | 0.28356 | 0.06301 | 0.00776 | 0.00000 |
| 32 | 0.64693 | 0.26588 | 0.07140 | 0.01579 | 0.00000 | 0.60866 | 0.25361 | 0.11587 | 0.02187 | 0.00000 |
| 33 | 0.78449 | 0.14234 | 0.06038 | 0.01279 | 0.00000 | 0.64821 | 0.24051 | 0.10205 | 0.00923 | 0.00000 |
| 34 | 0.69715 | 0.19226 | 0.09613 | 0.01446 | 0.00000 | 0.62577 | 0.27115 | 0.09356 | 0.00952 | 0.00000 |
| 35 | 0.66071 | 0.26696 | 0.06339 | 0.00894 | 0.00000 | 0.65078 | 0.22320 | 0.08715 | 0.03887 | 0.00000 |
| 36 | 0.69882 | 0.19904 | 0.07158 | 0.03055 | 0.00000 | 0.59450 | 0.22508 | 0.13517 | 0.04526 | 0.00000 |
| 37 | 0.76033 | 0.14601 | 0.07301 | 0.02065 | 0.00000 | 0.50560 | 0.32192 | 0.11521 | 0.05727 | 0.00000 |
| 38 | 0.72142 | 0.19619 | 0.06952 | 0.01287 | 0.00000 | 0.53934 | 0.25488 | 0.16678 | 0.03900 | 0.00000 |
| 39 | 0.67979 | 0.23744 | 0.03881 | 0.04395 | 0.00000 | 0.44469 | 0.30310 | 0.19174 | 0.04056 | 0.01991 |
| 40 | 0.68339 | 0.16129 | 0.10992 | 0.03883 | 0.00657 | 0.55141 | 0.28944 | 0.12183 | 0.03732 | 0.00000 |
| 41 | 0.72445 | 0.15207 | 0.09428 | 0.02920 | 0.00000 | 0.46688 | 0.26656 | 0.16640 | 0.10016 | 0.00000 |
| 42 | 0.62258 | 0.19572 | 0.10488 | 0.07682 | 0.00000 | 0.52720 | 0.28368 | 0.13556 | 0.02678 | 0.02678 |
| 43 | 0.60524 | 0.23234 | 0.13401 | 0.02840 | 0.00000 | 0.48270 | 0.22981 | 0.19521 | 0.08075 | 0.01154 |
| 44 | 0.59522 | 0.21509 | 0.15683 | 0.03286 | 0.00000 | 0.51150 | 0.21067 | 0.16651 | 0.08924 | 0.02208 |
| 45 | 0.59843 | 0.17073 | 0.13676 | 0.07666 | 0.01742 | 0.52174 | 0.31919 | 0.07211 | 0.04348 | 0.04348 |
| 46 | 0.53484 | 0.28950 | 0.10500 | 0.06183 | 0.00883 | 0.60278 | 0.23535 | 0.07349 | 0.07349 | 0.01490 |
| 47 | 0.49950 | 0.25521 | 0.19166 | 0.04270 | 0.01092 | 0.49039 | 0.23559 | 0.13751 | 0.11729 | 0.01921 |
| 48 | 0.60208 | 0.17292 | 0.16354 | 0.06146 | 0.00000 | 0.37076 | 0.32097 | 0.18538 | 0.09852 | 0.02436 |
| 49 | 0.55496 | 0.21875 | 0.10884 | 0.09052 | 0.02694 | 0.45793 | 0.28767 | 0.15264 | 0.10176 | 0.00000 |
| 50 | 0.57638 | 0.16360 | 0.15385 | 0.09642 | 0.00975 | 0.55217 | 0.14961 | 0.17913 | 0.08957 | 0.02953 |
| 51 | 0.57158 | 0.23860 | 0.14316 | 0.02333 | 0.02333 | 0.46636 | 0.23318 | 0.16682 | 0.11705 | 0.01659 |
| 52 | 0.55767 | 0.24233 | 0.12593 | 0.04233 | 0.03175 | 0.47163 | 0.18605 | 0.21395 | 0.11442 | 0.01395 |
| 53 | 0.51420 | 0.23344 | 0.12618 | 0.10726 | 0.01893 | 0.35236 | 0.28170 | 0.18297 | 0.12681 | 0.05616 |
| 54 | 0.47527 | 0.20753 | 0.17097 | 0.10968 | 0.03656 | 0.35197 | 0.30981 | 0.22548 | 0.08433 | 0.02841 |
| 55 | 0.51613 | 0.22581 | 0.20430 | 0.05376 | 0.00000 | 0.42844 | 0.20019 | 0.20019 | 0.15716 | 0.01403 |
| 56 | 0.42070 | 0.27079 | 0.17795 | 0.11219 | 0.01838 | 0.32194 | 0.22032 | 0.23742 | 0.20323 | 0.01709 |
| 57 | 0.42500 | 0.24500 | 0.21300 | 0.09600 | 0.02100 | 0.42110 | 0.26331 | 0.19297 | 0.10551 | 0.01711 |
| 58 | 0.52090 | 0.20257 | 0.13076 | 0.10182 | 0.04394 | 0.27003 | 0.36499 | 0.22948 | 0.09496 | 0.04055 |
| 59 | 0.45483 | 0.21561 | 0.19302 | 0.10267 | 0.03388 | 0.41481 | 0.22837 | 0.21409 | 0.07136 | 0.07136 |
| 60 | 0.36134 | 0.27733 | 0.22874 | 0.09615 | 0.03644 | 0.40971 | 0.29458 | 0.21332 | 0.03273 | 0.04966 |
| 61 | 0.45540 | 0.22246 | 0.22246 | 0.07765 | 0.02204 | 0.35005 | 0.29990 | 0.17503 | 0.11259 | 0.06244 |
| 62 | 0.41793 | 0.26892 | 0.16414 | 0.10477 | 0.04424 | 0.33374 | 0.30340 | 0.16626 | 0.13592 | 0.06068 |
| 63 | 0.45936 | 0.28325 | 0.13547 | 0.08128 | 0.04064 | 0.45952 | 0.17976 | 0.24048 | 0.10000 | 0.02024 |
| 64 | 0.48243 | 0.22432 | 0.13784 | 0.08649 | 0.06892 | 0.25847 | 0.32246 | 0.19322 | 0.12923 | 0.09661 |
| 65 | 0.32690 | 0.27284 | 0.27284 | 0.10940 | 0.01802 | 0.30529 | 0.32212 | 0.15264 | 0.16947 | 0.05048 |
| 66 | 0.37265 | 0.27078 | 0.27078 | 0.06836 | 0.01743 | 0.35119 | 0.20982 | 0.20982 | 0.19345 | 0.03571 |
| 67 | 0.29569 | 0.24071 | 0.20357 | 0.18574 | 0.07429 | 0.18678 | 0.22975 | 0.27107 | 0.24959 | 0.06281 |
| 68 | 0.31325 | 0.34423 | 0.20310 | 0.06196 | 0.07745 | 0.26792 | 0.23208 | 0.21502 | 0.26792 | 0.01706 |
| 69 | 0.33874 | 0.15135 | 0.18919 | 0.16937 | 0.15135 | 0.31694 | 0.24408 | 0.31694 | 0.12204 | 0.00000 |
| 70 | 0.46392 | 0.14605 | 0.21993 | 0.14605 | 0.02405 | 0.31624 | 0.21709 | 0.18291 | 0.16752 | 0.11624 |
| 71 | 0.36704 | 0.14607 | 0.19476 | 0.17041 | 0.12172 | 0.20450 | 0.13701 | 0.31697 | 0.22699 | 0.11452 |
| 72 | 0.23622 | 0.21850 | 0.27362 | 0.18110 | 0.09055 | 0.25000 | 0.21460 | 0.28540 | 0.10619 | 0.14381 |
| 73 | 0.35510 | 0.17755 | 0.24490 | 0.15510 | 0.06735 | 0.21759 | 0.29861 | 0.13426 | 0.16667 | 0.18287 |
| 74 | 0.25051 | 0.25051 | 0.22382 | 0.12526 | 0.14990 | 0.24190 | 0.25935 | 0.20698 | 0.18953 | 0.10224 |
| 75 | 0.30485 | 0.18707 | 0.23788 | 0.15242 | 0.11778 | 0.22748 | 0.11486 | 0.36712 | 0.16441 | 0.12613 |
| 76 | 0.32667 | 0.19333 | 0.15333 | 0.17333 | 0.15333 | 0.16068 | 0.13319 | 0.16068 | 0.26638 | 0.27907 |
| 77 | 0.26939 | 0.07755 | 0.15306 | 0.30816 | 0.19184 | 0.11217 | 0.15036 | 0.26492 | 0.22673 | 0.24582 |

| | | | | | | | | | | |
|---|---|---|---|---|---|---|---|---|---|---|
| 78 | 0.30874 | 0.16667 | 0.14208 | 0.21585 | 0.16667 | 0.00000 | 0.28099 | 0.25895 | 0.30028 | 0.15978 |
| 79 | 0.17183 | 0.22817 | 0.14366 | 0.20000 | 0.25634 | 0.08202 | 0.18297 | 0.16404 | 0.38801 | 0.18297 |
| 80 | 0.11304 | 0.28696 | 0.20000 | 0.14203 | 0.25797 | 0.16398 | 0.16398 | 0.20430 | 0.20430 | 0.26344 |
| 81 | 0.04902 | 0.28431 | 0.09477 | 0.33333 | 0.23856 | 0.05243 | 0.20974 | 0.23596 | 0.18352 | 0.31835 |
| 82 | 0.11034 | 0.14828 | 0.25862 | 0.18621 | 0.29655 | 0.12552 | 0.07531 | 0.17573 | 0.22594 | 0.39749 |
| 83 | 0.19167 | 0.15417 | 0.15417 | 0.11667 | 0.38333 | 0.05078 | 0.10156 | 0.25781 | 0.23047 | 0.35938 |
| 84 | 0.05474 | 0.18978 | 0.18978 | 0.27007 | 0.29562 | 0.06667 | 0.26667 | 0.20000 | 0.20000 | 0.26667 |
| 85 | 0.29213 | 0.05993 | 0.05993 | 0.05993 | 0.52809 | 0.09633 | 0.16055 | 0.19266 | 0.19266 | 0.35780 |
| 86 | 0.12500 | 0.06250 | 0.25000 | 0.18750 | 0.37500 | 0.00000 | 0.04478 | 0.27239 | 0.31716 | 0.36567 |
| 87 | 0.00000 | 0.12360 | 0.25094 | 0.18727 | 0.43820 | 0.14667 | 0.07556 | 0.03556 | 0.33333 | 0.40889 |
| 88 | 0.04329 | 0.12987 | 0.08658 | 0.17316 | 0.56710 | 0.00000 | 0.06736 | 0.16580 | 0.23316 | 0.53368 |
| 89 | 0.08182 | 0.04091 | 0.15909 | 0.31818 | 0.40000 | 0.07216 | 0.14433 | 0.10825 | 0.28351 | 0.39175 |
| 90+ | 0.00000 | 0.08723 | 0.21703 | 0.13084 | 0.56490 | 0.06683 | 0.13366 | 0.06683 | 0.46659 | 0.26610 |

*SM.1.4.3: For g = South Gloucestershire*

| Age | **Births** | | | | |
|---|---|---|---|---|---|
| | **CS1** | **CS2** | **CS3** | **CS4** | **CS5** |
| 17 | 0.87817 | 0.10066 | 0.02013 | 0.00104 | 0 |

| Age | **Immigrations** | | | | | **Emigrations** | | | | |
|---|---|---|---|---|---|---|---|---|---|---|
| | **CS1** | **CS2** | **CS3** | **CS4** | **CS5** | **CS1** | **CS2** | **CS3** | **CS4** | **CS5** |
| 17 | 0.91796 | 0.08204 | 0.00000 | 0.00000 | 0.00000 | 0.85382 | 0.12048 | 0.01526 | 0.01044 | 0.00000 |
| 18 | 0.84289 | 0.12420 | 0.03291 | 0.00000 | 0.00000 | 0.84199 | 0.12641 | 0.03160 | 0.00000 | 0.00000 |
| 19 | 0.83146 | 0.15691 | 0.01162 | 0.00000 | 0.00000 | 0.76547 | 0.19023 | 0.03989 | 0.00440 | 0.00000 |
| 20 | 0.77968 | 0.18508 | 0.03082 | 0.00442 | 0.00000 | 0.73893 | 0.22776 | 0.03331 | 0.00000 | 0.00000 |
| 21 | 0.74388 | 0.21067 | 0.03717 | 0.00828 | 0.00000 | 0.71805 | 0.20212 | 0.07451 | 0.00531 | 0.00000 |
| 22 | 0.74829 | 0.20473 | 0.04364 | 0.00333 | 0.00000 | 0.67715 | 0.24015 | 0.07086 | 0.01184 | 0.00000 |
| 23 | 0.76849 | 0.18719 | 0.03949 | 0.00484 | 0.00000 | 0.68622 | 0.24089 | 0.06933 | 0.00356 | 0.00000 |
| 24 | 0.73324 | 0.22192 | 0.04232 | 0.00252 | 0.00000 | 0.67169 | 0.26997 | 0.04902 | 0.00622 | 0.00311 |
| 25 | 0.74942 | 0.20439 | 0.04619 | 0.00000 | 0.00000 | 0.71181 | 0.22771 | 0.04600 | 0.01448 | 0.00000 |
| 26 | 0.74493 | 0.20356 | 0.04630 | 0.00521 | 0.00000 | 0.65068 | 0.27816 | 0.06219 | 0.00897 | 0.00000 |
| 27 | 0.76102 | 0.18346 | 0.05123 | 0.00429 | 0.00000 | 0.64039 | 0.27553 | 0.07389 | 0.01018 | 0.00000 |
| 28 | 0.75055 | 0.18858 | 0.05648 | 0.00439 | 0.00000 | 0.67398 | 0.23962 | 0.06952 | 0.01687 | 0.00000 |
| 29 | 0.69974 | 0.23576 | 0.05959 | 0.00491 | 0.00000 | 0.69472 | 0.22441 | 0.06849 | 0.01239 | 0.00000 |
| 30 | 0.77434 | 0.16909 | 0.04994 | 0.00664 | 0.00000 | 0.67538 | 0.24280 | 0.07008 | 0.01174 | 0.00000 |
| 31 | 0.78421 | 0.16072 | 0.05023 | 0.00485 | 0.00000 | 0.68178 | 0.20907 | 0.09952 | 0.00963 | 0.00000 |
| 32 | 0.73465 | 0.20180 | 0.05171 | 0.00934 | 0.00251 | 0.66561 | 0.25363 | 0.07350 | 0.00726 | 0.00000 |
| 33 | 0.75596 | 0.18968 | 0.05162 | 0.00274 | 0.00000 | 0.64548 | 0.26601 | 0.06650 | 0.01565 | 0.00636 |
| 34 | 0.77174 | 0.17947 | 0.04370 | 0.00509 | 0.00000 | 0.64442 | 0.25000 | 0.09512 | 0.01046 | 0.00000 |
| 35 | 0.78399 | 0.14804 | 0.05866 | 0.00931 | 0.00000 | 0.67705 | 0.23333 | 0.06612 | 0.02350 | 0.00000 |
| 36 | 0.78522 | 0.14532 | 0.06650 | 0.00296 | 0.00000 | 0.69512 | 0.19315 | 0.09433 | 0.01291 | 0.00449 |
| 37 | 0.71452 | 0.20961 | 0.06550 | 0.01037 | 0.00000 | 0.67455 | 0.23721 | 0.06969 | 0.01407 | 0.00448 |
| 38 | 0.73396 | 0.15951 | 0.08181 | 0.02119 | 0.00353 | 0.66004 | 0.24414 | 0.07168 | 0.02413 | 0.00000 |
| 39 | 0.71567 | 0.20920 | 0.05246 | 0.02267 | 0.00000 | 0.63798 | 0.24481 | 0.09570 | 0.02151 | 0.00000 |
| 40 | 0.74558 | 0.14374 | 0.08301 | 0.02306 | 0.00461 | 0.69853 | 0.19526 | 0.08252 | 0.02369 | 0.00000 |

| | | | | | | | | | | |
|---|---|---|---|---|---|---|---|---|---|---|
| 41 | 0.74242 | 0.16188 | 0.08054 | 0.01515 | 0.00000 | 0.65171 | 0.22801 | 0.08259 | 0.03770 | 0.00000 |
| 42 | 0.72702 | 0.19017 | 0.05369 | 0.02912 | 0.00000 | 0.56749 | 0.21863 | 0.17205 | 0.04183 | 0.00000 |
| 43 | 0.71443 | 0.15309 | 0.12169 | 0.01079 | 0.00000 | 0.63190 | 0.21966 | 0.11334 | 0.03511 | 0.00000 |
| 44 | 0.69007 | 0.15446 | 0.14243 | 0.01304 | 0.00000 | 0.54824 | 0.28118 | 0.10471 | 0.06588 | 0.00000 |
| 45 | 0.70322 | 0.12872 | 0.13588 | 0.02622 | 0.00596 | 0.52163 | 0.30662 | 0.10687 | 0.06489 | 0.00000 |
| 46 | 0.68826 | 0.21188 | 0.07962 | 0.01350 | 0.00675 | 0.64134 | 0.21669 | 0.08219 | 0.04483 | 0.01494 |
| 47 | 0.72612 | 0.14888 | 0.09129 | 0.03371 | 0.00000 | 0.57576 | 0.29201 | 0.08815 | 0.04408 | 0.00000 |
| 48 | 0.69679 | 0.13120 | 0.14723 | 0.02478 | 0.00000 | 0.53662 | 0.28873 | 0.08310 | 0.06620 | 0.02535 |
| 49 | 0.65714 | 0.19683 | 0.10794 | 0.01905 | 0.01905 | 0.55292 | 0.24095 | 0.14345 | 0.06267 | 0.00000 |
| 50 | 0.61008 | 0.21176 | 0.12773 | 0.05042 | 0.00000 | 0.54065 | 0.21274 | 0.13008 | 0.07588 | 0.04065 |
| 51 | 0.65928 | 0.19761 | 0.08518 | 0.04770 | 0.01022 | 0.50066 | 0.27785 | 0.16645 | 0.04718 | 0.00786 |
| 52 | 0.62887 | 0.16151 | 0.13402 | 0.07560 | 0.00000 | 0.47273 | 0.27552 | 0.18322 | 0.05315 | 0.01538 |
| 53 | 0.63802 | 0.19008 | 0.11405 | 0.04793 | 0.00992 | 0.37247 | 0.24747 | 0.26389 | 0.09975 | 0.01641 |
| 54 | 0.60579 | 0.22785 | 0.08861 | 0.06872 | 0.00904 | 0.46742 | 0.28822 | 0.16541 | 0.05013 | 0.02882 |
| 55 | 0.61726 | 0.19137 | 0.08818 | 0.07317 | 0.03002 | 0.49935 | 0.25419 | 0.19355 | 0.04516 | 0.00774 |
| 56 | 0.61538 | 0.22065 | 0.08704 | 0.05466 | 0.02227 | 0.42273 | 0.31545 | 0.18774 | 0.06003 | 0.01405 |
| 57 | 0.58700 | 0.18449 | 0.15304 | 0.06499 | 0.01048 | 0.47727 | 0.30909 | 0.10303 | 0.06364 | 0.04697 |
| 58 | 0.60300 | 0.21674 | 0.12446 | 0.04506 | 0.01073 | 0.48196 | 0.23665 | 0.21356 | 0.03752 | 0.03030 |
| 59 | 0.50630 | 0.23950 | 0.17437 | 0.06723 | 0.01261 | 0.42508 | 0.22630 | 0.23242 | 0.07492 | 0.04128 |
| 60 | 0.58314 | 0.30296 | 0.08884 | 0.02506 | 0.00000 | 0.49548 | 0.26355 | 0.14006 | 0.06928 | 0.03163 |
| 61 | 0.53846 | 0.15385 | 0.18510 | 0.07692 | 0.04567 | 0.35761 | 0.17314 | 0.37055 | 0.07443 | 0.02427 |
| 62 | 0.49091 | 0.17662 | 0.17662 | 0.07792 | 0.07792 | 0.33608 | 0.24547 | 0.17298 | 0.14498 | 0.10049 |
| 63 | 0.45556 | 0.23333 | 0.14167 | 0.11667 | 0.05278 | 0.34884 | 0.30444 | 0.17336 | 0.14165 | 0.03171 |
| 64 | 0.41515 | 0.26061 | 0.15455 | 0.10909 | 0.06061 | 0.32285 | 0.24319 | 0.27254 | 0.12159 | 0.03983 |
| 65 | 0.34675 | 0.26935 | 0.28793 | 0.05882 | 0.03715 | 0.29412 | 0.35294 | 0.18824 | 0.10588 | 0.05882 |
| 66 | 0.53659 | 0.26829 | 0.16028 | 0.03484 | 0.00000 | 0.32258 | 0.20699 | 0.27419 | 0.14785 | 0.04839 |
| 67 | 0.43494 | 0.13011 | 0.28253 | 0.10781 | 0.04461 | 0.20323 | 0.34516 | 0.23871 | 0.15484 | 0.05806 |
| 68 | 0.36752 | 0.26496 | 0.16239 | 0.14530 | 0.05983 | 0.24046 | 0.26718 | 0.29008 | 0.12595 | 0.07634 |
| 69 | 0.33484 | 0.28054 | 0.15385 | 0.20362 | 0.02715 | 0.28794 | 0.31128 | 0.18288 | 0.15175 | 0.06615 |
| 70 | 0.44726 | 0.23629 | 0.08439 | 0.10549 | 0.12658 | 0.22530 | 0.23715 | 0.30040 | 0.16206 | 0.07510 |
| 71 | 0.46512 | 0.21395 | 0.17674 | 0.03721 | 0.10698 | 0.22388 | 0.24876 | 0.23383 | 0.20896 | 0.08458 |
| 72 | 0.34211 | 0.10526 | 0.28947 | 0.15789 | 0.10526 | 0.28261 | 0.25543 | 0.24457 | 0.14130 | 0.07609 |
| 73 | 0.51563 | 0.16146 | 0.13542 | 0.05208 | 0.13542 | 0.19905 | 0.31754 | 0.23223 | 0.13270 | 0.11848 |
| 74 | 0.27174 | 0.25000 | 0.12500 | 0.15217 | 0.20109 | 0.20430 | 0.22581 | 0.31720 | 0.15054 | 0.10215 |
| 75 | 0.49749 | 0.15075 | 0.07538 | 0.07538 | 0.20101 | 0.18041 | 0.34536 | 0.21649 | 0.12887 | 0.12887 |
| 76 | 0.34483 | 0.14286 | 0.11330 | 0.11330 | 0.28571 | 0.18067 | 0.23529 | 0.20168 | 0.20168 | 0.18067 |
| 77 | 0.27488 | 0.15166 | 0.15166 | 0.22275 | 0.19905 | 0.17751 | 0.15976 | 0.23669 | 0.23669 | 0.18935 |
| 78 | 0.25000 | 0.27500 | 0.17500 | 0.12500 | 0.17500 | 0.14724 | 0.15951 | 0.25767 | 0.30061 | 0.13497 |
| 79 | 0.47879 | 0.12121 | 0.07879 | 0.12121 | 0.20000 | 0.07576 | 0.17677 | 0.37374 | 0.20707 | 0.16667 |
| 80 | 0.14198 | 0.19136 | 0.19136 | 0.16667 | 0.30864 | 0.05696 | 0.24051 | 0.25949 | 0.24051 | 0.20253 |
| 81 | 0.31013 | 0.07595 | 0.11392 | 0.15190 | 0.34810 | 0.08571 | 0.19286 | 0.25714 | 0.19286 | 0.27143 |
| 82 | 0.10638 | 0.21277 | 0.15603 | 0.26241 | 0.26241 | 0.12903 | 0.16935 | 0.23387 | 0.23387 | 0.23387 |
| 83 | 0.12069 | 0.00000 | 0.35345 | 0.23276 | 0.29310 | 0.09816 | 0.14110 | 0.14110 | 0.38037 | 0.23926 |
| 84 | 0.16794 | 0.16794 | 0.16794 | 0.27481 | 0.22137 | 0.09722 | 0.12500 | 0.20139 | 0.22222 | 0.35417 |
| 85 | 0.11024 | 0.08661 | 0.19685 | 0.16535 | 0.44094 | 0.04795 | 0.11644 | 0.17123 | 0.28767 | 0.37671 |
| 86 | 0.08621 | 0.04310 | 0.16379 | 0.20690 | 0.50000 | 0.09756 | 0.09756 | 0.09756 | 0.36585 | 0.34146 |
| 87 | 0.05738 | 0.22131 | 0.27869 | 0.22131 | 0.22131 | 0.04819 | 0.11446 | 0.16265 | 0.27711 | 0.39759 |

| | | | | | | | | | | |
|---|---|---|---|---|---|---|---|---|---|---|
| 88 | 0.12264 | 0.00000 | 0.12264 | 0.44340 | 0.31132 | 0.05405 | 0.20270 | 0.11486 | 0.14189 | 0.48649 |
| 89 | 0.13725 | 0.06863 | 0.33333 | 0.19608 | 0.26471 | 0.00000 | 0.02703 | 0.27027 | 0.27027 | 0.43243 |
| 90+ | 0.00000 | 0.07692 | 0.00000 | 0.38462 | 0.53846 | 0.00000 | 0.11186 | 0.14746 | 0.29661 | 0.44407 |

***SM.1.5: Mean annual healthcare resource costs per person, $\bar{\tau}_{s,\bar{a},c}$***

Note that Total is the sum over all considered healthcare settings and not just Community, Maternity, Mental Health and Acute NEL. Values are Pounds Sterling (£).

| Age group | Core Segment | Community | Maternity | Mental Health | Acute NEL | Total |
|---|---|---|---|---|---|---|
| 17-29 | CS1 | 12.3 | 49.3 | 11.9 | 35.3 | 288.4 |
| 17-29 | CS2 | 28.9 | 97.9 | 166.1 | 84.4 | 807.7 |
| 17-29 | CS3 | 65.1 | 108 | 678.3 | 242 | 1901.4 |
| 17-29 | CS4 | 349.1 | 108.3 | 1291.5 | 784.4 | 4501.1 |
| 17-29 | CS5 | 2786.9 | 0 | 1079.9 | 3860.1 | 10225.6 |
| 30-39 | CS1 | 18.9 | 122.5 | 4.6 | 35.1 | 416.7 |
| 30-39 | CS2 | 30.7 | 162.9 | 100.3 | 80.5 | 817.4 |
| 30-39 | CS3 | 80.6 | 157.1 | 414.8 | 208.7 | 1658.3 |
| 30-39 | CS4 | 202.2 | 170.7 | 1204.8 | 678.2 | 3895 |
| 30-39 | CS5 | 422.9 | 35.8 | 1435.6 | 1505.8 | 5839.2 |
| 40-49 | CS1 | 12.8 | 19.2 | 4.1 | 30.7 | 253.7 |
| 40-49 | CS2 | 22.9 | 26.5 | 68 | 66.6 | 548.9 |
| 40-49 | CS3 | 79 | 17 | 266.5 | 180.3 | 1264.5 |
| 40-49 | CS4 | 200.2 | 18 | 490.4 | 551.5 | 2652.4 |
| 40-49 | CS5 | 667.8 | 42.3 | 965.8 | 1820.7 | 5805.9 |
| 50-59 | CS1 | 15.8 | 0.5 | 0.8 | 34.7 | 268.9 |
| 50-59 | CS2 | 27.7 | 0 | 44.5 | 89.5 | 555.8 |
| 50-59 | CS3 | 96.6 | 0.2 | 148.8 | 188 | 1152.6 |
| 50-59 | CS4 | 315.6 | 0 | 272.7 | 516.6 | 2463.8 |
| 50-59 | CS5 | 888.3 | 0.5 | 528.3 | 1468.4 | 4971 |
| 60-69 | CS1 | 22.6 | 0 | 3 | 47 | 355.3 |
| 60-69 | CS2 | 38.7 | 0 | 27.6 | 101.7 | 648 |
| 60-69 | CS3 | 118.7 | 0 | 66.5 | 208.3 | 1182.7 |
| 60-69 | CS4 | 330.6 | 0 | 202 | 499.4 | 2361.4 |
| 60-69 | CS5 | 905.1 | 0 | 438.3 | 1359.8 | 4722.7 |
| 70-79 | CS1 | 37.8 | 0 | 3.5 | 79.8 | 551.7 |
| 70-79 | CS2 | 63.8 | 0 | 20.4 | 139.8 | 844.3 |
| 70-79 | CS3 | 182.9 | 0 | 44 | 280.2 | 1434.8 |
| 70-79 | CS4 | 437.7 | 0 | 144.7 | 544.5 | 2524.6 |
| 70-79 | CS5 | 1216.4 | 0.1 | 266.7 | 1575.1 | 5044.7 |
| 80-89 | CS1 | 124 | 0 | 7.7 | 128.8 | 791.3 |
| 80-89 | CS2 | 127.8 | 0 | 46.8 | 213.7 | 1125.5 |
| 80-89 | CS3 | 368.3 | 0 | 47.5 | 462 | 1895 |
| 80-89 | CS4 | 699.3 | 0 | 72.2 | 703.9 | 2865.6 |
| 80-89 | CS5 | 1813 | 0 | 242.8 | 1584.9 | 5347.5 |
| 90+ | CS1 | 526.3 | 0 | 47.1 | 386.6 | 1544.2 |
| 90+ | CS2 | 522.2 | 0 | 12 | 350.7 | 1514 |

| | | | | | | |
|---|---|---|---|---|---|---|
| 90+ | CS3 | 1028.9 | 0 | 45.7 | 671.9 | 2627.8 |
| 90+ | CS4 | 1691.1 | 0 | 44.9 | 1071.7 | 3974.9 |
| 90+ | CS5 | 2297.1 | 0 | 227.7 | 1722.9 | 5591.6 |

**Section SM.2: Tabulated versions of model results**

***SM.2.1: Annual population size, $n_{y,g,a,c}$ (Figure 3 of paper), under the 'Baseline upper' scenario***

*SM2.1.1: For g = Bristol*

| Year | State Name | 17-29 | 30-39 | 40-49 | 50-59 | 60-69 | 70-79 | 80-89 | 90+ |
|---|---|---|---|---|---|---|---|---|---|
| 2025 | CS1 | 106118 | 56667 | 35364 | 22420 | 12561 | 6435 | 2163 | 523 |
| 2025 | CS2 | 22454 | 22354 | 15925 | 12934 | 10316 | 6377 | 2276 | 348 |
| 2025 | CS3 | 4727 | 7169 | 7717 | 8429 | 9037 | 6586 | 3011 | 556 |
| 2025 | CS4 | 546 | 1491 | 2443 | 4215 | 5691 | 5224 | 3303 | 683 |
| 2025 | CS5 | 18 | 95 | 362 | 1107 | 2196 | 3253 | 3291 | 998 |
| 2026 | CS1 | 107667 | 57487 | 35748 | 22057 | 12747 | 6192 | 2138 | 461 |
| 2026 | CS2 | 22859 | 22199 | 16354 | 12885 | 10537 | 6540 | 2419 | 386 |
| 2026 | CS3 | 4734 | 7072 | 7864 | 8339 | 9150 | 6728 | 3123 | 582 |
| 2026 | CS4 | 541 | 1499 | 2511 | 4088 | 5710 | 5317 | 3315 | 694 |
| 2026 | CS5 | 18 | 98 | 357 | 1067 | 2205 | 3209 | 3245 | 1006 |
| 2027 | CS1 | 109032 | 58720 | 36164 | 21680 | 12753 | 5891 | 2292 | 445 |
| 2027 | CS2 | 23186 | 22176 | 16748 | 12800 | 10749 | 6544 | 2671 | 437 |
| 2027 | CS3 | 4778 | 6996 | 8032 | 8260 | 9214 | 6692 | 3330 | 644 |
| 2027 | CS4 | 535 | 1505 | 2581 | 4009 | 5736 | 5251 | 3436 | 722 |
| 2027 | CS5 | 17 | 98 | 357 | 1030 | 2224 | 3140 | 3261 | 1004 |
| 2028 | CS1 | 110569 | 59760 | 36541 | 21399 | 12785 | 5759 | 2280 | 415 |
| 2028 | CS2 | 23557 | 22097 | 17206 | 12796 | 10831 | 6621 | 2851 | 483 |
| 2028 | CS3 | 4807 | 6951 | 8216 | 8221 | 9196 | 6710 | 3527 | 683 |
| 2028 | CS4 | 529 | 1494 | 2651 | 3968 | 5705 | 5236 | 3566 | 754 |
| 2028 | CS5 | 16 | 96 | 365 | 1010 | 2207 | 3088 | 3300 | 1029 |
| 2029 | CS1 | 111777 | 60993 | 36942 | 21449 | 12708 | 5677 | 2205 | 420 |
| 2029 | CS2 | 23982 | 22136 | 17478 | 12880 | 10901 | 6720 | 2961 | 520 |
| 2029 | CS3 | 4867 | 6941 | 8323 | 8205 | 9217 | 6765 | 3645 | 721 |
| 2029 | CS4 | 530 | 1498 | 2695 | 3899 | 5727 | 5254 | 3647 | 790 |
| 2029 | CS5 | 16 | 95 | 369 | 984 | 2211 | 3065 | 3331 | 1055 |
| 2030 | CS1 | 113123 | 61982 | 37246 | 21664 | 12616 | 5582 | 2208 | 394 |
| 2030 | CS2 | 24388 | 22192 | 17651 | 13089 | 10956 | 6802 | 3053 | 544 |
| 2030 | CS3 | 4921 | 6969 | 8369 | 8281 | 9214 | 6817 | 3741 | 743 |
| 2030 | CS4 | 528 | 1511 | 2716 | 3890 | 5725 | 5271 | 3728 | 809 |
| 2030 | CS5 | 15 | 96 | 372 | 967 | 2218 | 3041 | 3372 | 1077 |
| 2031 | CS1 | 114416 | 62664 | 37742 | 21652 | 12578 | 5600 | 2175 | 360 |
| 2031 | CS2 | 24816 | 22200 | 17855 | 13223 | 11020 | 6910 | 3130 | 549 |
| 2031 | CS3 | 5008 | 6962 | 8443 | 8308 | 9203 | 6919 | 3831 | 742 |
| 2031 | CS4 | 537 | 1512 | 2740 | 3883 | 5695 | 5340 | 3806 | 807 |
| 2031 | CS5 | 15 | 97 | 377 | 954 | 2208 | 3053 | 3425 | 1072 |
| 2032 | CS1 | 115294 | 63410 | 38195 | 21863 | 12421 | 5577 | 2139 | 359 |
| 2032 | CS2 | 25251 | 22196 | 18091 | 13356 | 10983 | 7065 | 3158 | 564 |
| 2032 | CS3 | 5118 | 6934 | 8544 | 8350 | 9145 | 7064 | 3858 | 757 |
| 2032 | CS4 | 552 | 1504 | 2776 | 3883 | 5649 | 5447 | 3833 | 821 |
| 2032 | CS5 | 15 | 97 | 382 | 947 | 2186 | 3097 | 3450 | 1086 |

| | | | | | | | | | |
|---|---|---|---|---|---|---|---|---|---|
| 2033 | CS1 | 115576 | 64551 | 38588 | 22069 | 12233 | 5582 | 2103 | 364 |
| 2033 | CS2 | 25580 | 22295 | 18263 | 13531 | 10908 | 7183 | 3195 | 594 |
| 2033 | CS3 | 5229 | 6906 | 8625 | 8442 | 9054 | 7181 | 3886 | 792 |
| 2033 | CS4 | 573 | 1484 | 2804 | 3910 | 5579 | 5547 | 3853 | 858 |
| 2033 | CS5 | 16 | 95 | 386 | 945 | 2151 | 3153 | 3454 | 1128 |
| 2034 | CS1 | 115545 | 65489 | 39237 | 22229 | 12061 | 5608 | 2062 | 363 |
| 2034 | CS2 | 25818 | 22413 | 18405 | 13762 | 10822 | 7314 | 3211 | 613 |
| 2034 | CS3 | 5320 | 6899 | 8678 | 8576 | 8948 | 7301 | 3909 | 813 |
| 2034 | CS4 | 591 | 1471 | 2814 | 3969 | 5497 | 5640 | 3887 | 880 |
| 2034 | CS5 | 17 | 93 | 386 | 951 | 2109 | 3197 | 3484 | 1161 |
| 2035 | CS1 | 115477 | 66210 | 39766 | 22502 | 11926 | 5662 | 2009 | 359 |
| 2035 | CS2 | 26003 | 22596 | 18462 | 14034 | 10707 | 7470 | 3209 | 633 |
| 2035 | CS3 | 5395 | 6921 | 8689 | 8743 | 8826 | 7444 | 3911 | 833 |
| 2035 | CS4 | 608 | 1465 | 2809 | 4045 | 5400 | 5753 | 3901 | 902 |
| 2035 | CS5 | 17 | 92 | 384 | 964 | 2061 | 3259 | 3501 | 1192 |
| 2036 | CS1 | 115465 | 66827 | 40201 | 22798 | 11760 | 5759 | 1969 | 366 |
| 2036 | CS2 | 26098 | 22862 | 18513 | 14305 | 10596 | 7600 | 3210 | 654 |
| 2036 | CS3 | 5423 | 7002 | 8700 | 8910 | 8713 | 7556 | 3919 | 854 |
| 2036 | CS4 | 612 | 1478 | 2806 | 4129 | 5317 | 5830 | 3922 | 919 |
| 2036 | CS5 | 17 | 92 | 384 | 983 | 2022 | 3300 | 3524 | 1214 |
| 2037 | CS1 | 115012 | 67385 | 40821 | 23121 | 11587 | 5799 | 1928 | 412 |
| 2037 | CS2 | 26100 | 23092 | 18654 | 14583 | 10477 | 7687 | 3182 | 729 |
| 2037 | CS3 | 5436 | 7071 | 8740 | 9091 | 8605 | 7624 | 3867 | 950 |
| 2037 | CS4 | 616 | 1489 | 2809 | 4224 | 5246 | 5881 | 3864 | 1015 |
| 2037 | CS5 | 17 | 92 | 384 | 1008 | 1990 | 3335 | 3463 | 1319 |
| 2038 | CS1 | 114098 | 68353 | 41292 | 23441 | 11462 | 5828 | 1914 | 423 |
| 2038 | CS2 | 25918 | 23472 | 18736 | 14881 | 10409 | 7723 | 3198 | 766 |
| 2038 | CS3 | 5401 | 7177 | 8759 | 9283 | 8538 | 7642 | 3873 | 999 |
| 2038 | CS4 | 612 | 1505 | 2803 | 4325 | 5205 | 5887 | 3864 | 1070 |
| 2038 | CS5 | 17 | 92 | 382 | 1034 | 1972 | 3338 | 3447 | 1394 |
| 2039 | CS1 | 113170 | 69045 | 41833 | 23718 | 11493 | 5817 | 1911 | 423 |
| 2039 | CS2 | 25735 | 23779 | 18872 | 15075 | 10440 | 7740 | 3222 | 783 |
| 2039 | CS3 | 5368 | 7271 | 8804 | 9403 | 8532 | 7659 | 3894 | 1017 |
| 2039 | CS4 | 609 | 1523 | 2808 | 4387 | 5184 | 5907 | 3882 | 1092 |
| 2039 | CS5 | 17 | 93 | 382 | 1051 | 1953 | 3355 | 3455 | 1431 |
| 2040 | CS1 | 112058 | 69885 | 42231 | 23935 | 11612 | 5792 | 1910 | 426 |
| 2040 | CS2 | 25486 | 24126 | 18987 | 15202 | 10563 | 7739 | 3240 | 796 |
| 2040 | CS3 | 5316 | 7374 | 8852 | 9474 | 8604 | 7656 | 3911 | 1033 |
| 2040 | CS4 | 604 | 1542 | 2819 | 4419 | 5215 | 5911 | 3897 | 1109 |
| 2040 | CS5 | 17 | 94 | 383 | 1059 | 1954 | 3364 | 3464 | 1459 |
| 2041 | CS1 | 110897 | 70880 | 42458 | 24238 | 11629 | 5792 | 1934 | 421 |
| 2041 | CS2 | 25201 | 24542 | 19033 | 15378 | 10611 | 7753 | 3288 | 799 |
| 2041 | CS3 | 5255 | 7509 | 8869 | 9576 | 8623 | 7659 | 3965 | 1034 |
| 2041 | CS4 | 596 | 1571 | 2820 | 4464 | 5220 | 5907 | 3949 | 1111 |
| 2041 | CS5 | 17 | 95 | 383 | 1071 | 1950 | 3359 | 3505 | 1470 |
| 2042 | CS1 | 109652 | 71771 | 42738 | 24521 | 11728 | 5732 | 1953 | 417 |
| 2042 | CS2 | 24886 | 24952 | 19087 | 15561 | 10701 | 7699 | 3341 | 796 |

| | | | | | | | | | |
|---|---|---|---|---|---|---|---|---|---|
| 2042 | CS3 | 5186 | 7651 | 8883 | 9691 | 8683 | 7607 | 4029 | 1026 |
| 2042 | CS4 | 587 | 1606 | 2818 | 4521 | 5246 | 5870 | 4017 | 1101 |
| 2042 | CS5 | 17 | 98 | 382 | 1086 | 1953 | 3339 | 3566 | 1464 |
| 2043 | CS1 | 108587 | 72266 | 43236 | 24750 | 11829 | 5656 | 1975 | 416 |
| 2043 | CS2 | 24614 | 25236 | 19228 | 15708 | 10815 | 7621 | 3389 | 798 |
| 2043 | CS3 | 5127 | 7764 | 8923 | 9786 | 8772 | 7528 | 4087 | 1027 |
| 2043 | CS4 | 580 | 1638 | 2819 | 4569 | 5296 | 5811 | 4080 | 1102 |
| 2043 | CS5 | 16 | 100 | 380 | 1099 | 1968 | 3306 | 3624 | 1467 |
| 2044 | CS1 | 107740 | 72489 | 43674 | 25095 | 11920 | 5578 | 2000 | 413 |
| 2044 | CS2 | 24399 | 25417 | 19362 | 15876 | 10950 | 7537 | 3438 | 796 |
| 2044 | CS3 | 5080 | 7845 | 8963 | 9883 | 8887 | 7441 | 4149 | 1024 |
| 2044 | CS4 | 574 | 1664 | 2822 | 4607 | 5369 | 5743 | 4146 | 1100 |
| 2044 | CS5 | 16 | 103 | 380 | 1106 | 1993 | 3266 | 3687 | 1470 |
| 2045 | CS1 | 106993 | 72732 | 44027 | 25361 | 12072 | 5512 | 2030 | 407 |
| 2045 | CS2 | 24196 | 25590 | 19505 | 15980 | 11127 | 7449 | 3497 | 789 |
| 2045 | CS3 | 5034 | 7921 | 9017 | 9937 | 9032 | 7348 | 4222 | 1014 |
| 2045 | CS4 | 567 | 1688 | 2834 | 4623 | 5462 | 5665 | 4224 | 1089 |
| 2045 | CS5 | 16 | 105 | 380 | 1108 | 2027 | 3217 | 3762 | 1462 |
| 2046 | CS1 | 106217 | 73039 | 44360 | 25576 | 12237 | 5439 | 2068 | 407 |
| 2046 | CS2 | 23971 | 25739 | 19693 | 16067 | 11309 | 7356 | 3553 | 788 |
| 2046 | CS3 | 4982 | 7976 | 9104 | 9983 | 9182 | 7252 | 4287 | 1012 |
| 2046 | CS4 | 560 | 1702 | 2860 | 4638 | 5559 | 5587 | 4289 | 1087 |
| 2046 | CS5 | 16 | 106 | 383 | 1111 | 2065 | 3170 | 3824 | 1463 |
| 2047 | CS1 | 105641 | 73038 | 44679 | 25891 | 12413 | 5362 | 2092 | 413 |
| 2047 | CS2 | 23804 | 25769 | 19861 | 16219 | 11501 | 7260 | 3586 | 797 |
| 2047 | CS3 | 4943 | 7996 | 9175 | 10064 | 9347 | 7155 | 4326 | 1024 |
| 2047 | CS4 | 555 | 1710 | 2881 | 4666 | 5667 | 5511 | 4329 | 1097 |
| 2047 | CS5 | 16 | 107 | 385 | 1116 | 2109 | 3126 | 3866 | 1475 |

*SM2.1.2: For g = North Somerset*

| **Year** | **State Name** | **17-29** | **30-39** | **40-49** | **50-59** | **60-69** | **70-79** | **80-89** | **90+** |
|---|---|---|---|---|---|---|---|---|---|
| 2025 | CS1 | 20947 | 18096 | 17262 | 15528 | 12236 | 8516 | 3688 | 588 |
| 2025 | CS2 | 5313 | 6296 | 6605 | 7414 | 6638 | 5332 | 2282 | 332 |
| 2025 | CS3 | 1295 | 2156 | 3174 | 4580 | 5350 | 5111 | 2802 | 534 |
| 2025 | CS4 | 200 | 542 | 1097 | 2220 | 3011 | 3818 | 2681 | 617 |
| 2025 | CS5 | 12 | 45 | 139 | 530 | 1119 | 2050 | 2525 | 916 |
| 2026 | CS1 | 21209 | 18021 | 17332 | 14859 | 11896 | 7877 | 3548 | 521 |
| 2026 | CS2 | 5502 | 6508 | 6888 | 7500 | 7078 | 5564 | 2514 | 388 |
| 2026 | CS3 | 1297 | 2196 | 3286 | 4620 | 5640 | 5296 | 3074 | 609 |
| 2026 | CS4 | 188 | 534 | 1106 | 2169 | 3138 | 3853 | 2841 | 673 |
| 2026 | CS5 | 12 | 43 | 138 | 516 | 1146 | 2067 | 2560 | 962 |
| 2027 | CS1 | 21521 | 17955 | 17505 | 14281 | 11603 | 7202 | 3483 | 515 |
| 2027 | CS2 | 5651 | 6735 | 7146 | 7570 | 7423 | 5566 | 2880 | 447 |
| 2027 | CS3 | 1291 | 2259 | 3388 | 4627 | 5934 | 5281 | 3417 | 684 |
| 2027 | CS4 | 184 | 530 | 1105 | 2156 | 3281 | 3773 | 3093 | 732 |

| | | | | | | | | | |
|---|---|---|---|---|---|---|---|---|---|
| 2027 | CS5 | 12 | 42 | 130 | 513 | 1170 | 2021 | 2694 | 1010 |
| 2028 | CS1 | 21754 | 17968 | 17795 | 13791 | 11222 | 6783 | 3326 | 482 |
| 2028 | CS2 | 5836 | 6863 | 7395 | 7647 | 7650 | 5667 | 3119 | 494 |
| 2028 | CS3 | 1321 | 2276 | 3499 | 4642 | 6135 | 5327 | 3668 | 752 |
| 2028 | CS4 | 183 | 525 | 1129 | 2124 | 3410 | 3755 | 3297 | 797 |
| 2028 | CS5 | 12 | 39 | 124 | 510 | 1203 | 2003 | 2815 | 1064 |
| 2029 | CS1 | 21924 | 18100 | 17829 | 13512 | 10999 | 6351 | 3159 | 487 |
| 2029 | CS2 | 5969 | 6998 | 7645 | 7656 | 7911 | 5723 | 3299 | 544 |
| 2029 | CS3 | 1333 | 2298 | 3608 | 4661 | 6292 | 5397 | 3845 | 815 |
| 2029 | CS4 | 182 | 518 | 1152 | 2121 | 3510 | 3778 | 3457 | 864 |
| 2029 | CS5 | 12 | 37 | 127 | 505 | 1231 | 2008 | 2918 | 1133 |
| 2030 | CS1 | 22019 | 18170 | 17872 | 13333 | 10743 | 6109 | 2987 | 437 |
| 2030 | CS2 | 6101 | 7097 | 7824 | 7763 | 8074 | 5832 | 3403 | 565 |
| 2030 | CS3 | 1348 | 2325 | 3685 | 4708 | 6424 | 5497 | 3959 | 845 |
| 2030 | CS4 | 181 | 522 | 1167 | 2120 | 3615 | 3846 | 3563 | 911 |
| 2030 | CS5 | 12 | 37 | 128 | 494 | 1274 | 2041 | 2998 | 1197 |
| 2031 | CS1 | 22109 | 18145 | 18046 | 13136 | 10502 | 5914 | 2837 | 407 |
| 2031 | CS2 | 6238 | 7148 | 8014 | 7805 | 8235 | 5935 | 3484 | 589 |
| 2031 | CS3 | 1373 | 2337 | 3751 | 4736 | 6557 | 5595 | 4040 | 873 |
| 2031 | CS4 | 184 | 523 | 1177 | 2122 | 3711 | 3929 | 3657 | 944 |
| 2031 | CS5 | 12 | 37 | 127 | 493 | 1302 | 2087 | 3085 | 1239 |
| 2032 | CS1 | 22290 | 18141 | 18221 | 13002 | 10197 | 5818 | 2656 | 389 |
| 2032 | CS2 | 6348 | 7214 | 8182 | 7882 | 8311 | 6087 | 3486 | 624 |
| 2032 | CS3 | 1390 | 2358 | 3817 | 4781 | 6613 | 5745 | 4061 | 912 |
| 2032 | CS4 | 183 | 525 | 1193 | 2134 | 3759 | 4053 | 3710 | 990 |
| 2032 | CS5 | 12 | 37 | 128 | 493 | 1317 | 2153 | 3157 | 1290 |
| 2033 | CS1 | 22531 | 18159 | 18387 | 12981 | 9874 | 5706 | 2516 | 370 |
| 2033 | CS2 | 6429 | 7308 | 8348 | 7982 | 8294 | 6247 | 3486 | 654 |
| 2033 | CS3 | 1404 | 2388 | 3876 | 4844 | 6612 | 5897 | 4071 | 953 |
| 2033 | CS4 | 184 | 531 | 1206 | 2153 | 3785 | 4184 | 3743 | 1041 |
| 2033 | CS5 | 12 | 37 | 128 | 494 | 1330 | 2221 | 3200 | 1358 |
| 2034 | CS1 | 22649 | 18220 | 18574 | 12975 | 9541 | 5649 | 2348 | 369 |
| 2034 | CS2 | 6477 | 7401 | 8525 | 8080 | 8257 | 6409 | 3444 | 693 |
| 2034 | CS3 | 1408 | 2412 | 3943 | 4915 | 6586 | 6062 | 4036 | 1004 |
| 2034 | CS4 | 182 | 532 | 1217 | 2186 | 3791 | 4325 | 3745 | 1103 |
| 2034 | CS5 | 12 | 36 | 126 | 498 | 1336 | 2294 | 3223 | 1440 |
| 2035 | CS1 | 22659 | 18323 | 18652 | 13027 | 9271 | 5593 | 2233 | 352 |
| 2035 | CS2 | 6516 | 7486 | 8671 | 8214 | 8169 | 6571 | 3401 | 723 |
| 2035 | CS3 | 1417 | 2435 | 4007 | 5000 | 6522 | 6231 | 3989 | 1046 |
| 2035 | CS4 | 184 | 534 | 1233 | 2228 | 3767 | 4478 | 3723 | 1160 |
| 2035 | CS5 | 12 | 36 | 126 | 506 | 1328 | 2381 | 3224 | 1522 |
| 2036 | CS1 | 22710 | 18470 | 18706 | 13168 | 8976 | 5546 | 2127 | 347 |
| 2036 | CS2 | 6557 | 7577 | 8775 | 8391 | 8046 | 6709 | 3365 | 750 |
| 2036 | CS3 | 1432 | 2461 | 4058 | 5107 | 6430 | 6373 | 3952 | 1083 |
| 2036 | CS4 | 186 | 539 | 1250 | 2280 | 3725 | 4612 | 3714 | 1207 |
| 2036 | CS5 | 12 | 37 | 128 | 518 | 1314 | 2461 | 3232 | 1588 |
| 2037 | CS1 | 22647 | 18588 | 18796 | 13360 | 8715 | 5503 | 2013 | 362 |

| | | | | | | | | | |
|---|---|---|---|---|---|---|---|---|---|
| 2037 | CS2 | 6578 | 7637 | 8900 | 8576 | 7931 | 6820 | 3279 | 822 |
| 2037 | CS3 | 1443 | 2474 | 4118 | 5216 | 6343 | 6500 | 3841 | 1182 |
| 2037 | CS4 | 189 | 541 | 1271 | 2329 | 3689 | 4737 | 3620 | 1321 |
| 2037 | CS5 | 12 | 37 | 130 | 528 | 1305 | 2540 | 3151 | 1731 |
| 2038 | CS1 | 22462 | 18789 | 18871 | 13613 | 8499 | 5413 | 1955 | 353 |
| 2038 | CS2 | 6536 | 7767 | 8971 | 8785 | 7844 | 6857 | 3266 | 850 |
| 2038 | CS3 | 1435 | 2517 | 4148 | 5344 | 6274 | 6558 | 3811 | 1223 |
| 2038 | CS4 | 188 | 551 | 1280 | 2390 | 3659 | 4816 | 3596 | 1376 |
| 2038 | CS5 | 12 | 37 | 131 | 542 | 1296 | 2599 | 3125 | 1819 |
| 2039 | CS1 | 22272 | 18933 | 19014 | 13725 | 8397 | 5370 | 1891 | 343 |
| 2039 | CS2 | 6496 | 7854 | 9065 | 8936 | 7788 | 6929 | 3241 | 866 |
| 2039 | CS3 | 1427 | 2544 | 4185 | 5440 | 6225 | 6633 | 3782 | 1246 |
| 2039 | CS4 | 187 | 556 | 1290 | 2439 | 3638 | 4895 | 3581 | 1408 |
| 2039 | CS5 | 12 | 38 | 132 | 554 | 1290 | 2652 | 3117 | 1877 |
| 2040 | CS1 | 21982 | 19154 | 19095 | 13821 | 8366 | 5305 | 1864 | 323 |
| 2040 | CS2 | 6426 | 7974 | 9128 | 9048 | 7803 | 6949 | 3255 | 857 |
| 2040 | CS3 | 1413 | 2582 | 4215 | 5508 | 6226 | 6668 | 3797 | 1234 |
| 2040 | CS4 | 186 | 565 | 1300 | 2473 | 3642 | 4951 | 3604 | 1403 |
| 2040 | CS5 | 12 | 38 | 133 | 563 | 1288 | 2699 | 3141 | 1895 |
| 2041 | CS1 | 21708 | 19434 | 19095 | 13988 | 8307 | 5250 | 1850 | 308 |
| 2041 | CS2 | 6345 | 8119 | 9146 | 9190 | 7782 | 6976 | 3282 | 850 |
| 2041 | CS3 | 1396 | 2631 | 4225 | 5590 | 6203 | 6706 | 3825 | 1223 |
| 2041 | CS4 | 183 | 576 | 1304 | 2509 | 3634 | 5003 | 3639 | 1395 |
| 2041 | CS5 | 11 | 39 | 133 | 570 | 1284 | 2739 | 3178 | 1903 |
| 2042 | CS1 | 21464 | 19722 | 19114 | 14147 | 8279 | 5154 | 1855 | 289 |
| 2042 | CS2 | 6262 | 8257 | 9178 | 9327 | 7796 | 6940 | 3337 | 829 |
| 2042 | CS3 | 1377 | 2677 | 4242 | 5673 | 6211 | 6682 | 3890 | 1197 |
| 2042 | CS4 | 181 | 586 | 1310 | 2547 | 3644 | 5007 | 3711 | 1372 |
| 2042 | CS5 | 11 | 40 | 134 | 578 | 1286 | 2752 | 3245 | 1892 |
| 2043 | CS1 | 21282 | 19917 | 19188 | 14284 | 8303 | 5036 | 1857 | 276 |
| 2043 | CS2 | 6194 | 8348 | 9242 | 9453 | 7850 | 6857 | 3392 | 816 |
| 2043 | CS3 | 1363 | 2707 | 4273 | 5749 | 6253 | 6616 | 3956 | 1180 |
| 2043 | CS4 | 179 | 593 | 1321 | 2583 | 3672 | 4980 | 3785 | 1356 |
| 2043 | CS5 | 11 | 40 | 135 | 586 | 1295 | 2748 | 3316 | 1884 |
| 2044 | CS1 | 21133 | 20046 | 19284 | 14441 | 8334 | 4907 | 1865 | 262 |
| 2044 | CS2 | 6138 | 8404 | 9308 | 9594 | 7914 | 6755 | 3450 | 798 |
| 2044 | CS3 | 1350 | 2726 | 4300 | 5835 | 6310 | 6530 | 4030 | 1158 |
| 2044 | CS4 | 178 | 597 | 1329 | 2621 | 3712 | 4935 | 3869 | 1335 |
| 2044 | CS5 | 11 | 40 | 136 | 594 | 1309 | 2733 | 3398 | 1871 |
| 2045 | CS1 | 20988 | 20176 | 19402 | 14523 | 8407 | 4798 | 1873 | 252 |
| 2045 | CS2 | 6079 | 8465 | 9378 | 9694 | 8011 | 6644 | 3507 | 783 |
| 2045 | CS3 | 1337 | 2748 | 4332 | 5899 | 6392 | 6430 | 4104 | 1140 |
| 2045 | CS4 | 176 | 603 | 1338 | 2654 | 3766 | 4870 | 3954 | 1316 |
| 2045 | CS5 | 11 | 41 | 136 | 602 | 1328 | 2703 | 3486 | 1856 |
| 2046 | CS1 | 20830 | 20310 | 19540 | 14582 | 8519 | 4675 | 1879 | 245 |
| 2046 | CS2 | 6010 | 8533 | 9458 | 9767 | 8147 | 6512 | 3553 | 775 |
| 2046 | CS3 | 1321 | 2773 | 4370 | 5949 | 6505 | 6308 | 4166 | 1130 |

| | | | | | | | | | |
|---|---|---|---|---|---|---|---|---|---|
| 2046 | CS4 | 174 | 610 | 1350 | 2682 | 3838 | 4789 | 4026 | 1307 |
| 2046 | CS5 | 11 | 41 | 138 | 609 | 1356 | 2665 | 3564 | 1851 |
| 2047 | CS1 | 20726 | 20346 | 19660 | 14679 | 8657 | 4565 | 1881 | 240 |
| 2047 | CS2 | 5962 | 8561 | 9518 | 9859 | 8300 | 6389 | 3584 | 769 |
| 2047 | CS3 | 1309 | 2787 | 4394 | 6009 | 6630 | 6194 | 4213 | 1123 |
| 2047 | CS4 | 172 | 614 | 1356 | 2714 | 3915 | 4713 | 4085 | 1300 |
| 2047 | CS5 | 11 | 42 | 138 | 618 | 1385 | 2627 | 3633 | 1849 |

*SM2.1.3: For g = South Gloucestershire*

| Year | State Name | 17-29 | 30-39 | 40-49 | 50-59 | 60-69 | 70-79 | 80-89 | 90+ |
|---|---|---|---|---|---|---|---|---|---|
| 2025 | CS1 | 41533 | 31064 | 23325 | 18220 | 12636 | 6812 | 2740 | 382 |
| 2025 | CS2 | 9204 | 10580 | 9642 | 10325 | 8768 | 6161 | 2522 | 337 |
| 2025 | CS3 | 1933 | 3355 | 4382 | 6131 | 6960 | 5868 | 3210 | 525 |
| 2025 | CS4 | 233 | 655 | 1264 | 2710 | 3593 | 4374 | 3288 | 742 |
| 2025 | CS5 | 9 | 48 | 140 | 509 | 1128 | 2284 | 2929 | 1028 |
| 2026 | CS1 | 42155 | 31348 | 23826 | 17620 | 12495 | 6594 | 2622 | 354 |
| 2026 | CS2 | 9633 | 10913 | 9950 | 10154 | 9223 | 6239 | 2703 | 378 |
| 2026 | CS3 | 1936 | 3436 | 4560 | 6048 | 7217 | 5962 | 3365 | 573 |
| 2026 | CS4 | 219 | 669 | 1308 | 2712 | 3785 | 4373 | 3319 | 797 |
| 2026 | CS5 | 7 | 44 | 157 | 532 | 1178 | 2311 | 2929 | 1071 |
| 2027 | CS1 | 42737 | 31667 | 24344 | 17032 | 12474 | 6159 | 2718 | 336 |
| 2027 | CS2 | 9994 | 11217 | 10288 | 10010 | 9555 | 6201 | 2979 | 423 |
| 2027 | CS3 | 1950 | 3527 | 4744 | 5976 | 7401 | 5925 | 3627 | 618 |
| 2027 | CS4 | 210 | 687 | 1342 | 2726 | 3944 | 4282 | 3479 | 849 |
| 2027 | CS5 | 6 | 42 | 165 | 554 | 1234 | 2281 | 3015 | 1096 |
| 2028 | CS1 | 43270 | 31898 | 24974 | 16678 | 12182 | 6025 | 2610 | 345 |
| 2028 | CS2 | 10347 | 11407 | 10706 | 9888 | 9764 | 6244 | 3160 | 466 |
| 2028 | CS3 | 1987 | 3589 | 4938 | 5924 | 7525 | 5912 | 3838 | 675 |
| 2028 | CS4 | 206 | 700 | 1410 | 2708 | 4097 | 4220 | 3625 | 899 |
| 2028 | CS5 | 5 | 42 | 180 | 566 | 1302 | 2257 | 3103 | 1125 |
| 2029 | CS1 | 43750 | 32246 | 25477 | 16428 | 12086 | 5889 | 2526 | 322 |
| 2029 | CS2 | 10670 | 11641 | 11043 | 9832 | 9912 | 6337 | 3252 | 508 |
| 2029 | CS3 | 2037 | 3635 | 5126 | 5890 | 7655 | 5976 | 3925 | 722 |
| 2029 | CS4 | 207 | 708 | 1458 | 2691 | 4240 | 4264 | 3693 | 945 |
| 2029 | CS5 | 5 | 42 | 189 | 570 | 1367 | 2279 | 3159 | 1166 |
| 2030 | CS1 | 44235 | 32603 | 25845 | 16388 | 11882 | 5805 | 2461 | 308 |
| 2030 | CS2 | 10977 | 11840 | 11349 | 9854 | 9984 | 6428 | 3342 | 534 |
| 2030 | CS3 | 2085 | 3685 | 5290 | 5906 | 7730 | 6048 | 4014 | 748 |
| 2030 | CS4 | 208 | 714 | 1511 | 2684 | 4361 | 4321 | 3759 | 978 |
| 2030 | CS5 | 4 | 41 | 197 | 566 | 1432 | 2316 | 3212 | 1200 |
| 2031 | CS1 | 44677 | 32829 | 26303 | 16327 | 11718 | 5741 | 2384 | 303 |
| 2031 | CS2 | 11273 | 11978 | 11671 | 9840 | 10069 | 6573 | 3371 | 548 |
| 2031 | CS3 | 2140 | 3717 | 5453 | 5898 | 7803 | 6174 | 4062 | 762 |
| 2031 | CS4 | 212 | 716 | 1563 | 2674 | 4453 | 4425 | 3816 | 991 |
| 2031 | CS5 | 4 | 41 | 204 | 566 | 1473 | 2382 | 3275 | 1209 |

| | | | | | | | | | |
|---|---|---|---|---|---|---|---|---|---|
| 2032 | CS1 | 45083 | 33022 | 26800 | 16460 | 11349 | 5798 | 2245 | 320 |
| 2032 | CS2 | 11552 | 12070 | 12035 | 9890 | 10011 | 6771 | 3352 | 578 |
| 2032 | CS3 | 2213 | 3734 | 5616 | 5949 | 7783 | 6350 | 4041 | 800 |
| 2032 | CS4 | 224 | 718 | 1613 | 2693 | 4501 | 4551 | 3828 | 1026 |
| 2032 | CS5 | 5 | 41 | 211 | 569 | 1516 | 2447 | 3311 | 1237 |
| 2033 | CS1 | 45326 | 33377 | 27204 | 16575 | 11051 | 5829 | 2183 | 306 |
| 2033 | CS2 | 11753 | 12213 | 12356 | 9997 | 9888 | 6958 | 3355 | 603 |
| 2033 | CS3 | 2276 | 3755 | 5772 | 6035 | 7706 | 6517 | 4038 | 834 |
| 2033 | CS4 | 235 | 715 | 1663 | 2737 | 4494 | 4694 | 3828 | 1076 |
| 2033 | CS5 | 5 | 40 | 218 | 580 | 1528 | 2532 | 3316 | 1297 |
| 2034 | CS1 | 45439 | 33701 | 27631 | 16830 | 10689 | 5886 | 2091 | 314 |
| 2034 | CS2 | 11911 | 12361 | 12629 | 10190 | 9700 | 7154 | 3338 | 640 |
| 2034 | CS3 | 2323 | 3786 | 5903 | 6164 | 7576 | 6709 | 4008 | 885 |
| 2034 | CS4 | 242 | 716 | 1702 | 2795 | 4461 | 4846 | 3813 | 1137 |
| 2034 | CS5 | 5 | 39 | 222 | 591 | 1536 | 2613 | 3314 | 1364 |
| 2035 | CS1 | 45605 | 34017 | 27926 | 17165 | 10398 | 5913 | 2019 | 300 |
| 2035 | CS2 | 12051 | 12503 | 12867 | 10427 | 9527 | 7324 | 3311 | 659 |
| 2035 | CS3 | 2371 | 3815 | 6021 | 6314 | 7450 | 6891 | 3976 | 908 |
| 2035 | CS4 | 251 | 716 | 1742 | 2863 | 4409 | 5014 | 3795 | 1177 |
| 2035 | CS5 | 6 | 39 | 228 | 604 | 1529 | 2719 | 3303 | 1419 |
| 2036 | CS1 | 45762 | 34317 | 28163 | 17519 | 10118 | 5922 | 1984 | 291 |
| 2036 | CS2 | 12143 | 12672 | 13072 | 10696 | 9322 | 7499 | 3292 | 673 |
| 2036 | CS3 | 2400 | 3862 | 6123 | 6487 | 7304 | 7062 | 3961 | 919 |
| 2036 | CS4 | 255 | 722 | 1775 | 2949 | 4344 | 5171 | 3789 | 1198 |
| 2036 | CS5 | 6 | 39 | 232 | 626 | 1517 | 2817 | 3305 | 1450 |
| 2037 | CS1 | 45651 | 34583 | 28468 | 17888 | 9848 | 5954 | 1914 | 316 |
| 2037 | CS2 | 12176 | 12823 | 13291 | 10981 | 9128 | 7643 | 3232 | 736 |
| 2037 | CS3 | 2416 | 3905 | 6231 | 6669 | 7164 | 7204 | 3878 | 1005 |
| 2037 | CS4 | 258 | 728 | 1811 | 3038 | 4284 | 5301 | 3713 | 1301 |
| 2037 | CS5 | 6 | 39 | 237 | 646 | 1508 | 2903 | 3236 | 1562 |
| 2038 | CS1 | 45385 | 35035 | 28669 | 18317 | 9683 | 5880 | 1907 | 311 |
| 2038 | CS2 | 12120 | 13061 | 13429 | 11319 | 8998 | 7678 | 3244 | 760 |
| 2038 | CS3 | 2407 | 3978 | 6300 | 6882 | 7067 | 7255 | 3874 | 1037 |
| 2038 | CS4 | 257 | 740 | 1835 | 3147 | 4236 | 5378 | 3700 | 1354 |
| 2038 | CS5 | 6 | 39 | 241 | 673 | 1497 | 2970 | 3213 | 1634 |
| 2039 | CS1 | 45094 | 35350 | 28937 | 18656 | 9601 | 5866 | 1896 | 301 |
| 2039 | CS2 | 12058 | 13245 | 13597 | 11586 | 8923 | 7729 | 3265 | 761 |
| 2039 | CS3 | 2398 | 4038 | 6375 | 7054 | 6999 | 7323 | 3892 | 1035 |
| 2039 | CS4 | 257 | 752 | 1859 | 3233 | 4200 | 5460 | 3722 | 1364 |
| 2039 | CS5 | 6 | 40 | 244 | 694 | 1485 | 3035 | 3232 | 1659 |
| 2040 | CS1 | 44705 | 35746 | 29156 | 18920 | 9627 | 5806 | 1901 | 294 |
| 2040 | CS2 | 11964 | 13454 | 13737 | 11812 | 8935 | 7722 | 3300 | 764 |
| 2040 | CS3 | 2381 | 4105 | 6440 | 7202 | 6994 | 7336 | 3927 | 1035 |
| 2040 | CS4 | 255 | 766 | 1879 | 3310 | 4197 | 5504 | 3757 | 1372 |
| 2040 | CS5 | 6 | 40 | 247 | 713 | 1481 | 3084 | 3261 | 1678 |
| 2041 | CS1 | 44311 | 36224 | 29273 | 19233 | 9622 | 5763 | 1911 | 285 |
| 2041 | CS2 | 11854 | 13697 | 13822 | 12063 | 8917 | 7729 | 3351 | 754 |

| | | | | | | | | | |
|---|---|---|---|---|---|---|---|---|---|
| 2041 | CS3 | 2359 | 4186 | 6481 | 7363 | 6967 | 7356 | 3987 | 1021 |
| 2041 | CS4 | 252 | 784 | 1891 | 3391 | 4180 | 5543 | 3822 | 1363 |
| 2041 | CS5 | 6 | 41 | 248 | 732 | 1473 | 3120 | 3320 | 1678 |
| 2042 | CS1 | 43908 | 36691 | 29380 | 19564 | 9696 | 5631 | 1947 | 271 |
| 2042 | CS2 | 11729 | 13941 | 13884 | 12335 | 8974 | 7625 | 3436 | 736 |
| 2042 | CS3 | 2335 | 4274 | 6510 | 7533 | 7009 | 7277 | 4085 | 994 |
| 2042 | CS4 | 249 | 805 | 1900 | 3478 | 4205 | 5515 | 3917 | 1338 |
| 2042 | CS5 | 6 | 43 | 250 | 753 | 1480 | 3126 | 3402 | 1661 |
| 2043 | CS1 | 43587 | 36984 | 29621 | 19824 | 9772 | 5515 | 1979 | 266 |
| 2043 | CS2 | 11624 | 14107 | 14005 | 12569 | 9057 | 7503 | 3516 | 729 |
| 2043 | CS3 | 2315 | 4338 | 6560 | 7686 | 7075 | 7172 | 4179 | 984 |
| 2043 | CS4 | 247 | 822 | 1913 | 3559 | 4251 | 5452 | 4014 | 1330 |
| 2043 | CS5 | 6 | 44 | 251 | 773 | 1497 | 3106 | 3489 | 1655 |
| 2044 | CS1 | 43351 | 37141 | 29849 | 20105 | 9913 | 5368 | 2013 | 259 |
| 2044 | CS2 | 11545 | 14217 | 14125 | 12788 | 9208 | 7336 | 3602 | 721 |
| 2044 | CS3 | 2300 | 4384 | 6609 | 7825 | 7195 | 7024 | 4284 | 974 |
| 2044 | CS4 | 245 | 836 | 1926 | 3630 | 4327 | 5359 | 4120 | 1322 |
| 2044 | CS5 | 6 | 45 | 253 | 790 | 1524 | 3067 | 3585 | 1649 |
| 2045 | CS1 | 43139 | 37308 | 30077 | 20296 | 10100 | 5245 | 2033 | 251 |
| 2045 | CS2 | 11465 | 14327 | 14244 | 12959 | 9398 | 7189 | 3662 | 710 |
| 2045 | CS3 | 2285 | 4431 | 6661 | 7937 | 7345 | 6890 | 4370 | 958 |
| 2045 | CS4 | 243 | 850 | 1940 | 3690 | 4419 | 5268 | 4218 | 1307 |
| 2045 | CS5 | 6 | 46 | 254 | 805 | 1556 | 3023 | 3683 | 1636 |
| 2046 | CS1 | 42897 | 37505 | 30302 | 20450 | 10298 | 5124 | 2049 | 249 |
| 2046 | CS2 | 11372 | 14429 | 14379 | 13104 | 9609 | 7031 | 3720 | 706 |
| 2046 | CS3 | 2265 | 4471 | 6724 | 8033 | 7516 | 6744 | 4449 | 953 |
| 2046 | CS4 | 241 | 860 | 1958 | 3743 | 4528 | 5165 | 4306 | 1302 |
| 2046 | CS5 | 6 | 47 | 256 | 818 | 1597 | 2970 | 3773 | 1631 |
| 2047 | CS1 | 42738 | 37549 | 30511 | 20657 | 10505 | 5010 | 2065 | 248 |
| 2047 | CS2 | 11307 | 14466 | 14502 | 13272 | 9832 | 6880 | 3766 | 705 |
| 2047 | CS3 | 2251 | 4490 | 6777 | 8141 | 7698 | 6604 | 4514 | 952 |
| 2047 | CS4 | 240 | 867 | 1974 | 3799 | 4645 | 5064 | 4378 | 1304 |
| 2047 | CS5 | 5 | 48 | 258 | 832 | 1640 | 2918 | 3847 | 1635 |

***SM.2.2: Indexed cost projections at selected healthcare settings, $r_{y,g,s,a,c}$ (Figure 4 of paper), under the 'Baseline upper' scenario***

| **Area** | **Year** | **Community** | **Maternity** | **Mental Health** | **Acute NEL** | **Total** |
|---|---|---|---|---|---|---|
| BNSSG | 2024 | 100 | 100 | 100 | 100 | 100 |
| BNSSG | 2025 | 101.5 | 101.7 | 101.1 | 101.3 | 101.4 |
| BNSSG | 2026 | 103 | 103 | 102.3 | 102.6 | 102.7 |
| BNSSG | 2027 | 105 | 104.4 | 103.5 | 104.2 | 104.1 |
| BNSSG | 2028 | 107.1 | 105.7 | 104.8 | 105.8 | 105.5 |
| BNSSG | 2029 | 109.2 | 107.1 | 106.2 | 107.4 | 107 |
| BNSSG | 2030 | 110.9 | 108.4 | 107.5 | 108.8 | 108.3 |
| BNSSG | 2031 | 112.4 | 109.5 | 108.8 | 110.2 | 109.6 |

| | | | | | | |
|---|---|---|---|---|---|---|
| BNSSG | 2032 | 114 | 110.5 | 110.2 | 111.6 | 110.8 |
| BNSSG | 2033 | 115.7 | 111.6 | 111.4 | 112.9 | 112 |
| BNSSG | 2034 | 117.4 | 112.5 | 112.5 | 114.1 | 113.1 |
| BNSSG | 2035 | 118.8 | 113.4 | 113.5 | 115.3 | 114.1 |
| BNSSG | 2036 | 120.1 | 114.2 | 114.5 | 116.3 | 115 |
| BNSSG | 2037 | 121.9 | 114.8 | 115.4 | 117.5 | 116 |
| BNSSG | 2038 | 123.2 | 115.6 | 116.1 | 118.4 | 116.8 |
| BNSSG | 2039 | 124.2 | 116.2 | 116.8 | 119.3 | 117.5 |
| BNSSG | 2040 | 125.1 | 116.8 | 117.4 | 120 | 118.2 |
| BNSSG | 2041 | 125.8 | 117.5 | 118 | 120.7 | 118.8 |
| BNSSG | 2042 | 126.4 | 118.1 | 118.6 | 121.3 | 119.4 |
| BNSSG | 2043 | 127.1 | 118.5 | 119.1 | 122 | 120 |
| BNSSG | 2044 | 127.8 | 118.7 | 119.7 | 122.6 | 120.5 |
| BNSSG | 2045 | 128.4 | 118.9 | 120.2 | 123.2 | 121.1 |
| BNSSG | 2046 | 129.1 | 119.1 | 120.7 | 123.8 | 121.6 |
| BNSSG | 2047 | 129.7 | 119.1 | 121.1 | 124.3 | 122.1 |
| Bristol | 2024 | 100 | 100 | 100 | 100 | 100 |
| Bristol | 2025 | 100 | 101.4 | 100.3 | 100.3 | 100.6 |
| Bristol | 2026 | 100.2 | 102.4 | 100.7 | 100.6 | 101.1 |
| Bristol | 2027 | 101 | 103.6 | 101.2 | 101.1 | 101.8 |
| Bristol | 2028 | 101.9 | 104.8 | 101.7 | 101.8 | 102.6 |
| Bristol | 2029 | 102.9 | 106.2 | 102.4 | 102.6 | 103.5 |
| Bristol | 2030 | 103.8 | 107.4 | 103.2 | 103.4 | 104.3 |
| Bristol | 2031 | 104.5 | 108.4 | 103.9 | 104.1 | 105.1 |
| Bristol | 2032 | 105.3 | 109.4 | 104.8 | 104.9 | 105.9 |
| Bristol | 2033 | 106.3 | 110.5 | 105.5 | 105.7 | 106.7 |
| Bristol | 2034 | 107.3 | 111.3 | 106.2 | 106.4 | 107.5 |
| Bristol | 2035 | 108.2 | 112.1 | 106.8 | 107.1 | 108.2 |
| Bristol | 2036 | 109 | 112.8 | 107.5 | 107.8 | 108.8 |
| Bristol | 2037 | 110.3 | 113.4 | 108 | 108.6 | 109.5 |
| Bristol | 2038 | 111.3 | 114.1 | 108.4 | 109.2 | 110.1 |
| Bristol | 2039 | 112 | 114.5 | 108.8 | 109.7 | 110.6 |
| Bristol | 2040 | 112.7 | 115 | 109.1 | 110.2 | 111.1 |
| Bristol | 2041 | 113.2 | 115.6 | 109.4 | 110.7 | 111.5 |
| Bristol | 2042 | 113.7 | 116.2 | 109.7 | 111.2 | 112 |
| Bristol | 2043 | 114.3 | 116.4 | 110.1 | 111.6 | 112.4 |
| Bristol | 2044 | 114.8 | 116.5 | 110.4 | 112 | 112.7 |
| Bristol | 2045 | 115.2 | 116.6 | 110.7 | 112.4 | 113.1 |
| Bristol | 2046 | 115.8 | 116.7 | 111 | 112.9 | 113.5 |
| Bristol | 2047 | 116.3 | 116.6 | 111.2 | 113.3 | 113.8 |
| NSom | 2024 | 100 | 100 | 100 | 100 | 100 |
| NSom | 2025 | 103 | 101.4 | 101.6 | 102.3 | 102.1 |
| NSom | 2026 | 105.9 | 102.6 | 103.4 | 104.7 | 104.2 |
| NSom | 2027 | 109.5 | 103.9 | 105.2 | 107.3 | 106.4 |
| NSom | 2028 | 112.9 | 105.2 | 107.2 | 109.9 | 108.6 |
| NSom | 2029 | 116.3 | 106.4 | 109 | 112.4 | 110.6 |
| NSom | 2030 | 119.1 | 107.3 | 110.8 | 114.6 | 112.5 |

| | | | | | | |
|---|---|---|---|---|---|---|
| NSom | 2031 | 121.6 | 108 | 112.5 | 116.6 | 114.2 |
| NSom | 2032 | 124.1 | 108.7 | 114.1 | 118.6 | 115.8 |
| NSom | 2033 | 126.5 | 109.6 | 115.6 | 120.4 | 117.3 |
| NSom | 2034 | 128.9 | 110.4 | 116.9 | 122.1 | 118.7 |
| NSom | 2035 | 131 | 111.1 | 118.2 | 123.7 | 119.9 |
| NSom | 2036 | 132.9 | 112 | 119.4 | 125.1 | 121.1 |
| NSom | 2037 | 135.3 | 112.6 | 120.5 | 126.6 | 122.2 |
| NSom | 2038 | 136.9 | 113.3 | 121.5 | 127.7 | 123.2 |
| NSom | 2039 | 138.2 | 113.8 | 122.3 | 128.7 | 124 |
| NSom | 2040 | 138.9 | 114.4 | 123 | 129.5 | 124.7 |
| NSom | 2041 | 139.7 | 115.2 | 123.7 | 130.2 | 125.3 |
| NSom | 2042 | 140.3 | 116 | 124.4 | 130.9 | 125.9 |
| NSom | 2043 | 141 | 116.5 | 125 | 131.6 | 126.5 |
| NSom | 2044 | 141.6 | 116.8 | 125.6 | 132.2 | 127.1 |
| NSom | 2045 | 142.2 | 117.2 | 126.3 | 132.8 | 127.6 |
| NSom | 2046 | 142.9 | 117.5 | 126.9 | 133.4 | 128.1 |
| NSom | 2047 | 143.5 | 117.6 | 127.4 | 134 | 128.6 |
| SGlos | 2024 | 100 | 100 | 100 | 100 | 100 |
| SGlos | 2025 | 102.1 | 102.4 | 102.2 | 102 | 101.9 |
| SGlos | 2026 | 104.2 | 104.4 | 104.4 | 104 | 103.8 |
| SGlos | 2027 | 106.7 | 106.3 | 106.6 | 106.1 | 105.8 |
| SGlos | 2028 | 109.2 | 107.9 | 108.9 | 108.2 | 107.6 |
| SGlos | 2029 | 111.4 | 109.7 | 111.1 | 110.2 | 109.5 |
| SGlos | 2030 | 113.5 | 111.3 | 113.2 | 112 | 111.2 |
| SGlos | 2031 | 115.3 | 112.7 | 115.1 | 113.8 | 112.8 |
| SGlos | 2032 | 117.1 | 113.9 | 117.2 | 115.5 | 114.3 |
| SGlos | 2033 | 119 | 115.2 | 119 | 117.1 | 115.8 |
| SGlos | 2034 | 121 | 116.4 | 120.7 | 118.8 | 117.2 |
| SGlos | 2035 | 122.6 | 117.5 | 122.3 | 120.2 | 118.6 |
| SGlos | 2036 | 124 | 118.5 | 123.8 | 121.5 | 119.8 |
| SGlos | 2037 | 126 | 119.3 | 125.1 | 123 | 121 |
| SGlos | 2038 | 127.6 | 120.3 | 126.3 | 124.2 | 122.1 |
| SGlos | 2039 | 128.7 | 121 | 127.4 | 125.3 | 123.1 |
| SGlos | 2040 | 129.8 | 121.8 | 128.5 | 126.3 | 124 |
| SGlos | 2041 | 130.7 | 122.6 | 129.4 | 127.3 | 124.9 |
| SGlos | 2042 | 131.5 | 123.5 | 130.4 | 128.2 | 125.8 |
| SGlos | 2043 | 132.5 | 124.1 | 131.4 | 129.1 | 126.6 |
| SGlos | 2044 | 133.4 | 124.4 | 132.3 | 130 | 127.4 |
| SGlos | 2045 | 134.2 | 124.8 | 133.2 | 130.9 | 128.1 |
| SGlos | 2046 | 135.1 | 125.1 | 134 | 131.7 | 128.9 |
| SGlos | 2047 | 136 | 125.2 | 134.7 | 132.5 | 129.6 |

***SM.2.3: Total cost growth to 2047 over different scenarios (Figure 5A of paper), under the 'Baseline upper' scenario***

| **Change to cost to deliver activity** | **Change to upwards Core Segment transition rate** | **Cost growth to 2047** |
|---|---|---|

| | | |
|---|---|---|
| -25.0% | -70.0% | -36.6 |
| -25.0% | -60.0% | -32.6 |
| -25.0% | -50.0% | -28.5 |
| -25.0% | -40.0% | -24.4 |
| -25.0% | -30.0% | -20.3 |
| -25.0% | -20.0% | -16.3 |
| -25.0% | -10.0% | -12.4 |
| -25.0% | 0.0% | -8.4 |
| -25.0% | 10.0% | -4.6 |
| -25.0% | 20.0% | -0.8 |
| -25.0% | 30.0% | 2.9 |
| -25.0% | 40.0% | 6.6 |
| -25.0% | 50.0% | 10.2 |
| -25.0% | 60.0% | 13.7 |
| -25.0% | 70.0% | 17.1 |
| -19.6% | -70.0% | -32.1 |
| -19.6% | -60.0% | -27.8 |
| -19.6% | -50.0% | -23.4 |
| -19.6% | -40.0% | -19 |
| -19.6% | -30.0% | -14.6 |
| -19.6% | -20.0% | -10.4 |
| -19.6% | -10.0% | -6.1 |
| -19.6% | 0.0% | -1.9 |
| -19.6% | 10.0% | 2.2 |
| -19.6% | 20.0% | 6.3 |
| -19.6% | 30.0% | 10.3 |
| -19.6% | 40.0% | 14.2 |
| -19.6% | 50.0% | 18 |
| -19.6% | 60.0% | 21.8 |
| -19.6% | 70.0% | 25.5 |
| -14.3% | -70.0% | -27.6 |
| -14.3% | -60.0% | -23 |
| -14.3% | -50.0% | -18.3 |
| -14.3% | -40.0% | -13.6 |
| -14.3% | -30.0% | -9 |
| -14.3% | -20.0% | -4.4 |
| -14.3% | -10.0% | 0.2 |
| -14.3% | 0.0% | 4.6 |
| -14.3% | 10.0% | 9 |
| -14.3% | 20.0% | 13.4 |
| -14.3% | 30.0% | 17.6 |
| -14.3% | 40.0% | 21.8 |
| -14.3% | 50.0% | 25.9 |
| -14.3% | 60.0% | 29.9 |
| -14.3% | 70.0% | 33.9 |
| -8.9% | -70.0% | -23.1 |
| -8.9% | -60.0% | -18.2 |

| | | |
|---|---|---|
| -8.9% | -50.0% | -13.2 |
| -8.9% | -40.0% | -8.2 |
| -8.9% | -30.0% | -3.3 |
| -8.9% | -20.0% | 1.6 |
| -8.9% | -10.0% | 6.4 |
| -8.9% | 0.0% | 11.2 |
| -8.9% | 10.0% | 15.9 |
| -8.9% | 20.0% | 20.4 |
| -8.9% | 30.0% | 25 |
| -8.9% | 40.0% | 29.4 |
| -8.9% | 50.0% | 33.8 |
| -8.9% | 60.0% | 38.1 |
| -8.9% | 70.0% | 42.2 |
| -3.6% | -70.0% | -18.5 |
| -3.6% | -60.0% | -13.4 |
| -3.6% | -50.0% | -8.1 |
| -3.6% | -40.0% | -2.8 |
| -3.6% | -30.0% | 2.4 |
| -3.6% | -20.0% | 7.6 |
| -3.6% | -10.0% | 12.7 |
| -3.6% | 0.0% | 17.7 |
| -3.6% | 10.0% | 22.7 |
| -3.6% | 20.0% | 27.5 |
| -3.6% | 30.0% | 32.3 |
| -3.6% | 40.0% | 37 |
| -3.6% | 50.0% | 41.6 |
| -3.6% | 60.0% | 46.2 |
| -3.6% | 70.0% | 50.6 |
| 1.8% | -70.0% | -14 |
| 1.8% | -60.0% | -8.6 |
| 1.8% | -50.0% | -3 |
| 1.8% | -40.0% | 2.6 |
| 1.8% | -30.0% | 8.1 |
| 1.8% | -20.0% | 13.5 |
| 1.8% | -10.0% | 18.9 |
| 1.8% | 0.0% | 24.3 |
| 1.8% | 10.0% | 29.5 |
| 1.8% | 20.0% | 34.6 |
| 1.8% | 30.0% | 39.7 |
| 1.8% | 40.0% | 44.6 |
| 1.8% | 50.0% | 49.5 |
| 1.8% | 60.0% | 54.3 |
| 1.8% | 70.0% | 59 |
| 7.1% | -70.0% | -9.5 |
| 7.1% | -60.0% | -3.7 |
| 7.1% | -50.0% | 2.1 |
| 7.1% | -40.0% | 8 |

| | | |
|---|---|---|
| 7.1% | -30.0% | 13.8 |
| 7.1% | -20.0% | 19.5 |
| 7.1% | -10.0% | 25.2 |
| 7.1% | 0.0% | 30.8 |
| 7.1% | 10.0% | 36.3 |
| 7.1% | 20.0% | 41.7 |
| 7.1% | 30.0% | 47 |
| 7.1% | 40.0% | 52.2 |
| 7.1% | 50.0% | 57.4 |
| 7.1% | 60.0% | 62.4 |
| 7.1% | 70.0% | 67.3 |
| 12.5% | -70.0% | -4.9 |
| 12.5% | -60.0% | 1.1 |
| 12.5% | -50.0% | 7.2 |
| 12.5% | -40.0% | 13.4 |
| 12.5% | -30.0% | 19.5 |
| 12.5% | -20.0% | 25.5 |
| 12.5% | -10.0% | 31.5 |
| 12.5% | 0.0% | 37.3 |
| 12.5% | 10.0% | 43.1 |
| 12.5% | 20.0% | 48.8 |
| 12.5% | 30.0% | 54.4 |
| 12.5% | 40.0% | 59.8 |
| 12.5% | 50.0% | 65.3 |
| 12.5% | 60.0% | 70.5 |
| 12.5% | 70.0% | 75.7 |
| 17.9% | -70.0% | -0.4 |
| 17.9% | -60.0% | 5.9 |
| 17.9% | -50.0% | 12.3 |
| 17.9% | -40.0% | 18.8 |
| 17.9% | -30.0% | 25.2 |
| 17.9% | -20.0% | 31.5 |
| 17.9% | -10.0% | 37.7 |
| 17.9% | 0.0% | 43.9 |
| 17.9% | 10.0% | 49.9 |
| 17.9% | 20.0% | 55.9 |
| 17.9% | 30.0% | 61.7 |
| 17.9% | 40.0% | 67.5 |
| 17.9% | 50.0% | 73.1 |
| 17.9% | 60.0% | 78.7 |
| 17.9% | 70.0% | 84.1 |
| 23.2% | -70.0% | 4.1 |
| 23.2% | -60.0% | 10.7 |
| 23.2% | -50.0% | 17.4 |
| 23.2% | -40.0% | 24.2 |
| 23.2% | -30.0% | 30.9 |
| 23.2% | -20.0% | 37.5 |

| | | |
|---|---|---|
| 23.2% | -10.0% | 44 |
| 23.2% | 0.0% | 50.4 |
| 23.2% | 10.0% | 56.7 |
| 23.2% | 20.0% | 63 |
| 23.2% | 30.0% | 69.1 |
| 23.2% | 40.0% | 75.1 |
| 23.2% | 50.0% | 81 |
| 23.2% | 60.0% | 86.8 |
| 23.2% | 70.0% | 92.4 |
| 28.6% | -70.0% | 8.6 |
| 28.6% | -60.0% | 15.5 |
| 28.6% | -50.0% | 22.5 |
| 28.6% | -40.0% | 29.6 |
| 28.6% | -30.0% | 36.6 |
| 28.6% | -20.0% | 43.4 |
| 28.6% | -10.0% | 50.3 |
| 28.6% | 0.0% | 56.9 |
| 28.6% | 10.0% | 63.6 |
| 28.6% | 20.0% | 70 |
| 28.6% | 30.0% | 76.4 |
| 28.6% | 40.0% | 82.7 |
| 28.6% | 50.0% | 88.9 |
| 28.6% | 60.0% | 94.9 |
| 28.6% | 70.0% | 100.8 |
| 33.9% | -70.0% | 13.2 |
| 33.9% | -60.0% | 20.3 |
| 33.9% | -50.0% | 27.6 |
| 33.9% | -40.0% | 35 |
| 33.9% | -30.0% | 42.2 |
| 33.9% | -20.0% | 49.4 |
| 33.9% | -10.0% | 56.5 |
| 33.9% | 0.0% | 63.5 |
| 33.9% | 10.0% | 70.3 |
| 33.9% | 20.0% | 77.1 |
| 33.9% | 30.0% | 83.8 |
| 33.9% | 40.0% | 90.3 |
| 33.9% | 50.0% | 96.7 |
| 33.9% | 60.0% | 103 |
| 33.9% | 70.0% | 109.2 |
| 39.3% | -70.0% | 17.7 |
| 39.3% | -60.0% | 25.1 |
| 39.3% | -50.0% | 32.7 |
| 39.3% | -40.0% | 40.4 |
| 39.3% | -30.0% | 47.9 |
| 39.3% | -20.0% | 55.4 |
| 39.3% | -10.0% | 62.8 |
| 39.3% | 0.0% | 70 |

| | | |
|---|---|---|
| 39.3% | 10.0% | 77.2 |
| 39.3% | 20.0% | 84.2 |
| 39.3% | 30.0% | 91.1 |
| 39.3% | 40.0% | 97.9 |
| 39.3% | 50.0% | 104.6 |
| 39.3% | 60.0% | 111.1 |
| 39.3% | 70.0% | 117.5 |
| 44.6% | -70.0% | 22.2 |
| 44.6% | -60.0% | 29.9 |
| 44.6% | -50.0% | 37.8 |
| 44.6% | -40.0% | 45.8 |
| 44.6% | -30.0% | 53.6 |
| 44.6% | -20.0% | 61.4 |
| 44.6% | -10.0% | 69 |
| 44.6% | 0.0% | 76.6 |
| 44.6% | 10.0% | 84 |
| 44.6% | 20.0% | 91.3 |
| 44.6% | 30.0% | 98.5 |
| 44.6% | 40.0% | 105.5 |
| 44.6% | 50.0% | 112.4 |
| 44.6% | 60.0% | 119.2 |
| 44.6% | 70.0% | 125.9 |
| 50.0% | -70.0% | 26.7 |
| 50.0% | -60.0% | 34.7 |
| 50.0% | -50.0% | 43 |
| 50.0% | -40.0% | 51.2 |
| 50.0% | -30.0% | 59.3 |
| 50.0% | -20.0% | 67.3 |
| 50.0% | -10.0% | 75.3 |
| 50.0% | 0.0% | 83.1 |
| 50.0% | 10.0% | 90.8 |
| 50.0% | 20.0% | 98.4 |
| 50.0% | 30.0% | 105.8 |
| 50.0% | 40.0% | 113.1 |
| 50.0% | 50.0% | 120.3 |
| 50.0% | 60.0% | 127.4 |
| 50.0% | 70.0% | 134.3 |

***SM.2.4: Annual cost growth to 2047 on comparison to year 2024 (Figure 5B and 5C of paper), under the 'Baseline upper' scenario***

| **Year** | **Scenario** | | | |
|---|---|---|---|---|
| | **Baseline** | **Efficiency** | **Prevention** | **Prevention & Efficiency** |
| 2024 | 0 | 0 | 0 | 0 |
| 2025 | 1.4 | -0.6 | 1.3 | -0.7 |
| 2026 | 2.7 | 0.6 | 2.5 | 0.5 |

| | | | | |
|---|---|---|---|---|
| 2027 | 4.1 | 2.1 | 3.8 | 1.7 |
| 2028 | 5.5 | 3.5 | 5 | 3 |
| 2029 | 7 | 4.9 | 6.3 | 4.2 |
| 2030 | 8.3 | 6.2 | 7.5 | 5.4 |
| 2031 | 9.6 | 7.4 | 8.7 | 6.5 |
| 2032 | 10.8 | 8.6 | 9.8 | 7.6 |
| 2033 | 12 | 9.7 | 10.8 | 8.6 |
| 2034 | 13.1 | 10.8 | 11.8 | 9.6 |
| 2035 | 14.1 | 11.8 | 12.8 | 10.5 |
| 2036 | 15 | 12.7 | 13.6 | 11.3 |
| 2037 | 16 | 13.6 | 14.5 | 12.1 |
| 2038 | 16.8 | 14.4 | 15.3 | 12.9 |
| 2039 | 17.5 | 15.1 | 15.9 | 13.5 |
| 2040 | 18.2 | 15.8 | 16.5 | 14.1 |
| 2041 | 18.8 | 16.4 | 17.1 | 14.7 |
| 2042 | 19.4 | 17 | 17.6 | 15.2 |
| 2043 | 20 | 17.5 | 18.2 | 15.7 |
| 2044 | 20.5 | 18.1 | 18.7 | 16.2 |
| 2045 | 21.1 | 18.6 | 19.1 | 16.6 |
| 2046 | 21.6 | 19.1 | 19.6 | 17.1 |
| 2047 | 22.1 | 19.6 | 20.1 | 17.6 |
